# COVID-19 Pandemic Analysis

**DOI:** 10.1101/2021.07.12.21260326

**Authors:** Victor A.J. van Lint

## Abstract

The development of coronavirus disease (COVID-19) deaths in selected nations and states is compared to the result of calculations using a conventional SEIR model of pandemic development. The model is based on the infection multiplier, *R*_*0*_, defined as the number of people infected by each infectious person. The infection rate increases exponentially when *R*_*0*_ >1.0; it remains constant at R_*0*_ = 1.0 and decreases for *R*_*0*_ < 1.0. *R*_*0*_ is determined by population behavior (frequency and proximity of interactions) and the ease by which a victim is infected by an infectious person (virulence of the virus). It is reduced by herd immunity when a large fraction of the population acquires immunity by vaccination or by recovering from infection.

The daily death rates in the U.S. and northern Europe exhibited peaks in April/May 2020 and Dec. 2020/Jan. 2021 with more a modest rate during the summer of 2020 and a gradually decreasing rate since Jan. 2021. The model produces this type of oscillatory response if it assumes that the population’s *R*_*0*_ responds to information reported about the pandemic, but with a delay between infections and resulting behavioral adjustments. Oscillatory behavior is typical of a control loop with delay in its feedback.

**The analysis concludes that:** Given the history of *R*_*0*_ the model predicts the development of pandemic deaths. However, since *R*_*0*_ is determined by the population’s behavior, control of the pandemic in democracies depends primarily on preparation and the persuasive power of political and scientific authorities. Data for S. Korea and New Zealand demonstrate the effectiveness of such methods.
For each death in the U.S. about 169 persons were infected, but fewer than half of them were identified as cases.
The pandemic was prolonged in the U.S. because the population chose to keep *R*_*0*_ near 1.0 by relaxing restrictions once the death rate subsided.
Initial values of *R*_*0*_ as high as 5.0 were observed, leading to infections doubling about every 2 days. If unabated, the resulting exponential growth increases the infected population by a factor of about 5000 before the death from the first infections is recorded.
Arrival from Italy probably initiated the pandemic in the eastern U.S., but, by the time the first death was recorded the number of domestic infections exceeded by far those that were imported. Import restrictions beyond this point are ineffective except in delaying the arrival of more virulent mutations.
If no social restrictions had been adopted, approximately 1.6 million deaths would have resulted in the U.S. The vaccine, although developed and deployed at record speed, was too late to ameliorate this result.
A third peak in death rate in Sept. 2021 may be prevented if more than 80% of the population is vaccinated.

## Introduction

The objective of this analysis is to produce a model to guide the response to future pandemic threats.

The coronavirus disease (COVID-19) pandemic stressed the United States and the world in general more severely than any event since World War II. As of May 31, 2021, more than 560,000 people died in the U.S. (1700 deaths per million inhabitants, dpM), and the peak death rates in some states had exceeded 10 dpM/day. This report analyzes data on accumulated deaths for various geographic regions since March 1, 2020, and compares the data to the calculations with the SEIR (Susceptible, Exposed, Infectious, Recovered) mathematical model presented in Appendix A.

In most western nations and U.S. states the pandemic outpaced initial attempts at control, progressed rapidly until about 0.1% of the population had died, and then continued to spread at a lower rate. This behavior is consistent with a model in which the public adjusts its average interaction rate in response to pandemic information, such as data on testing, hospitalizations and deaths.

We believe the most reliable data are the cumulative deaths attributed to the virus; hence, we define:

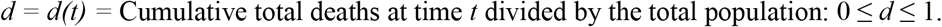

It is assumed that the effectiveness of treatment does not change during the pandemic, such that *d(t)* is proportional to the fraction of the population that were infected at an earlier time, most of whom have recovered. Because deaths must be reported, the primary uncertainty lies in the identified cause of death. Despite the numerous politically motivated attacks on the Internet, we are reasonably sure that these data are reliable. We have not relied on data for the reported number of “cases” because they depend on the intensity of testing, which has clearly changed during the pandemic. The disadvantage of using deaths is that they lag the infection events by about one month.

Our approach selects and analyzes published data on accumulated COVID-19 deaths during the period from March 1, 2020, through May 31, 2021, for representative nations and states. Fortunately, Wikipedia has maintained the appropriate data logs, presumably recording data reported by official government sources, such as the U.S. Center for Disease Control. The data are presented as deaths per million (dpM) population to provide a meaningful comparison of severity between areas of different populations.

## Results

A range of histories of the accumulation of COVID-19 deaths are shown in Figure 1.^*^

**Figure 1:**
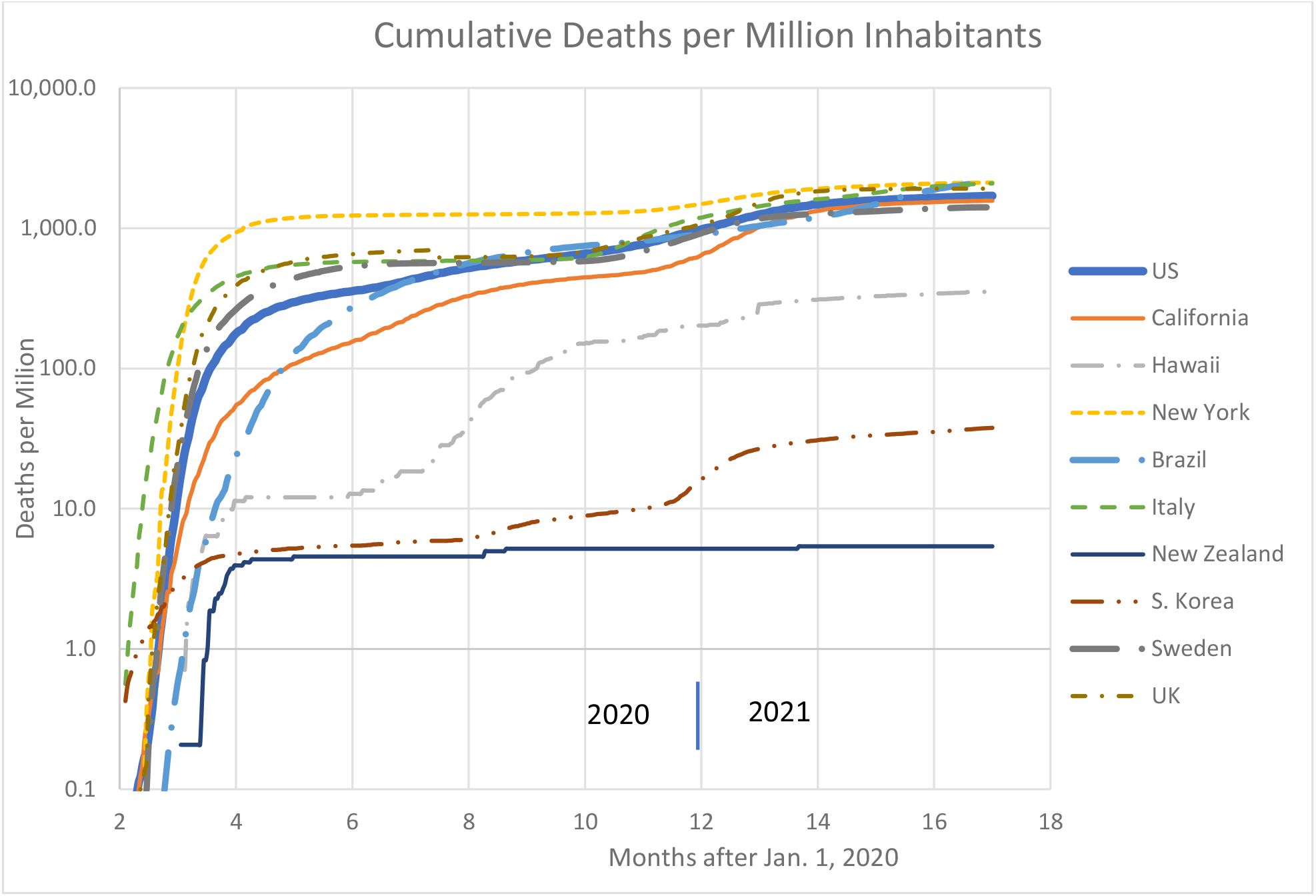
COVID-19 dpM: a range of histories.

The data all start with exponential growth. The growth rates (i.e., slopes on the semilogarithmic plots) gradually decrease, presumably as the population adjusts its behavior. The data appear to fall into three classes:

> Those that appear to have stopped the pandemic at an early stage and maintained control; examples include South Korea, New Zealand, Alaska and Hawaii, although Hawaii experienced a reemergence during the summer and early fall of 2020.
>
> Those that are continuing to experience an increase at a modest rate; California is an example.
>
> Those that appeared to level off in the summer of 2020 after an early dramatic increase; examples include New York, Italy, Sweden, Michigan, and the United Kingdom. Most of them resumed dramatic growth in the fall.

The U.S. is a composite of states at different stages of development.

It might appear from Fig. 1 that the pandemic development after the summer 2020 was mild compared to its March/April growths, but that is an illusion produced by the semilogarithmic plot. Figure 2 presents the daily deaths for the same nations/states as compared to the daily deaths during the 2017-18 annual influenza. It is clear that there were two waves with comparable intensities that are much more severe than the annual flu. Since daily data are inherently noisy and are distorted by reporting delays on the weekends, all daily data were calculated as averages over 7 days.

**Figure 2.**
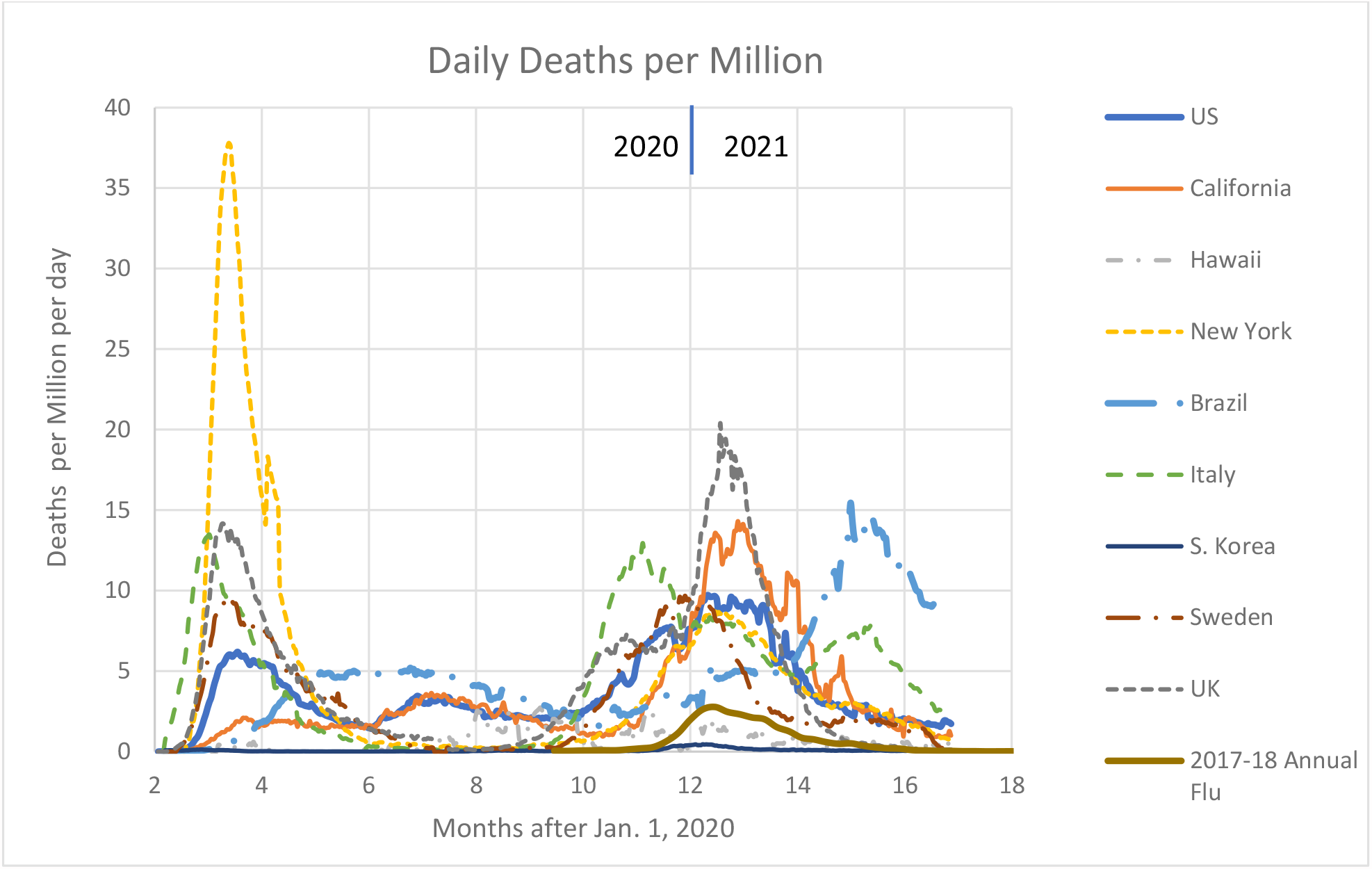
Daily deaths per million for same states/nations as Figure 1 (7-day averages)

In many areas (e.g., California, Brazil) the peak death rates in January 2021 exceeded those encountered during the earlier peak.

### Inception

The death rates soon after the onset of the pandemic in an area provides a measure of the threat. The evolution of the death rates in the earliest onsets is illustrated in Figure 3.

**Figure 3.**
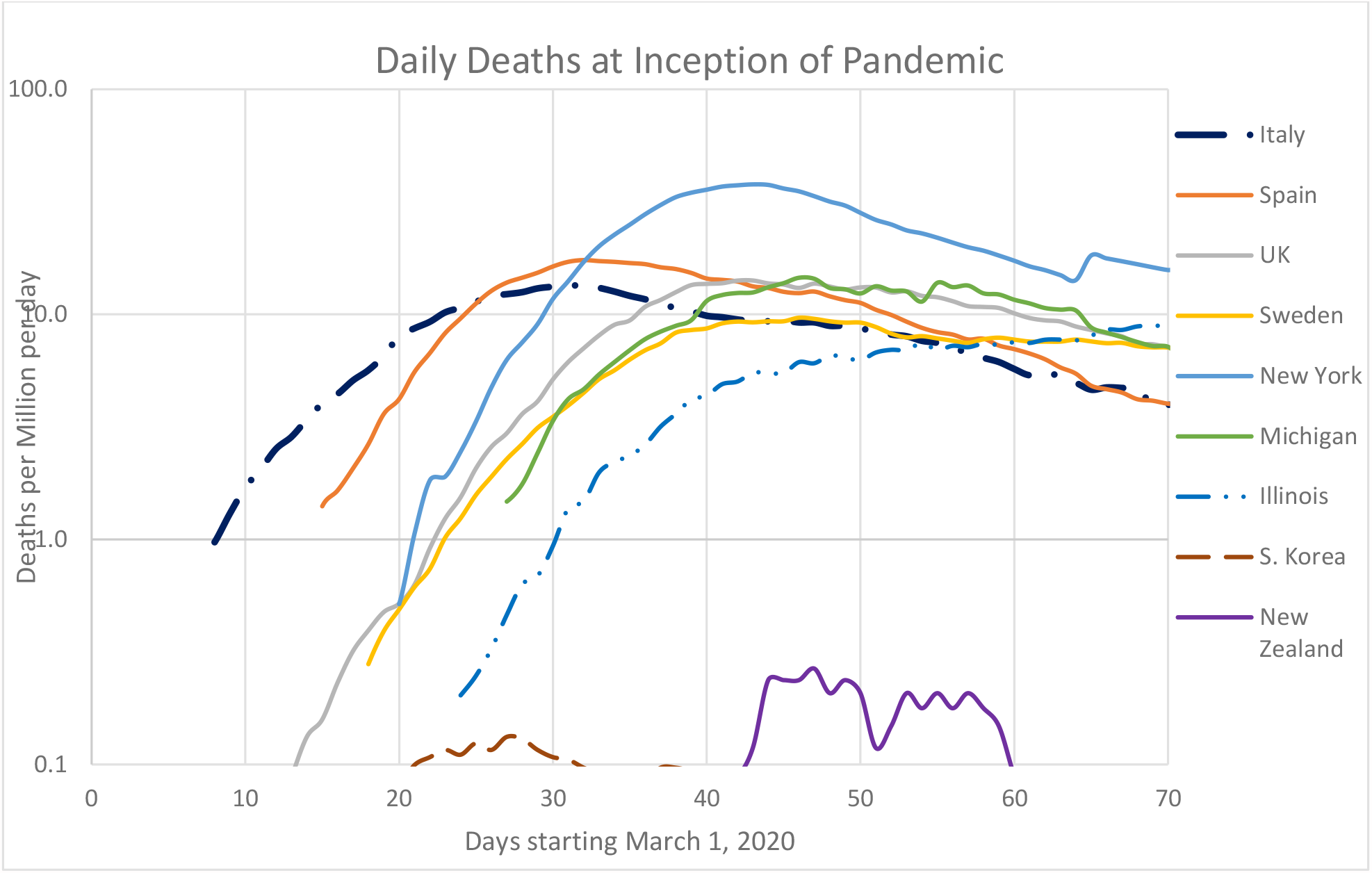
Daily deaths per million after the earlier inceptions (7-day averages)

The time sequence of onsets reflects the transmission of the infection from its initial injection into Europe in Italy to Spain, and then to UK and Sweden, and in parallel to New York, Michigan and Illinois. Even though S. Korea and New Zealand were nearest to the source of the pandemic in China, their readiness enabled them to delay and suppress infections.

The slopes of the semilogarithmic plots of death rates in Figure 3 measure the exponentiation times (1.44 times the doubling time) of the exponential growths, which depend on *R*_*0*_, the infection multiplier (i.e., the number of people infected by each infectious person), albeit the value of *R*_*0*_ at the time they were infected rather than the time they died. These slopes, again averaged over 7 days, are shown in Figure 4. These curves are noisy because they depend on differences between daily deaths that fluctuate due to reporting delays even when averaged over 7 days.

**Figure 4.**
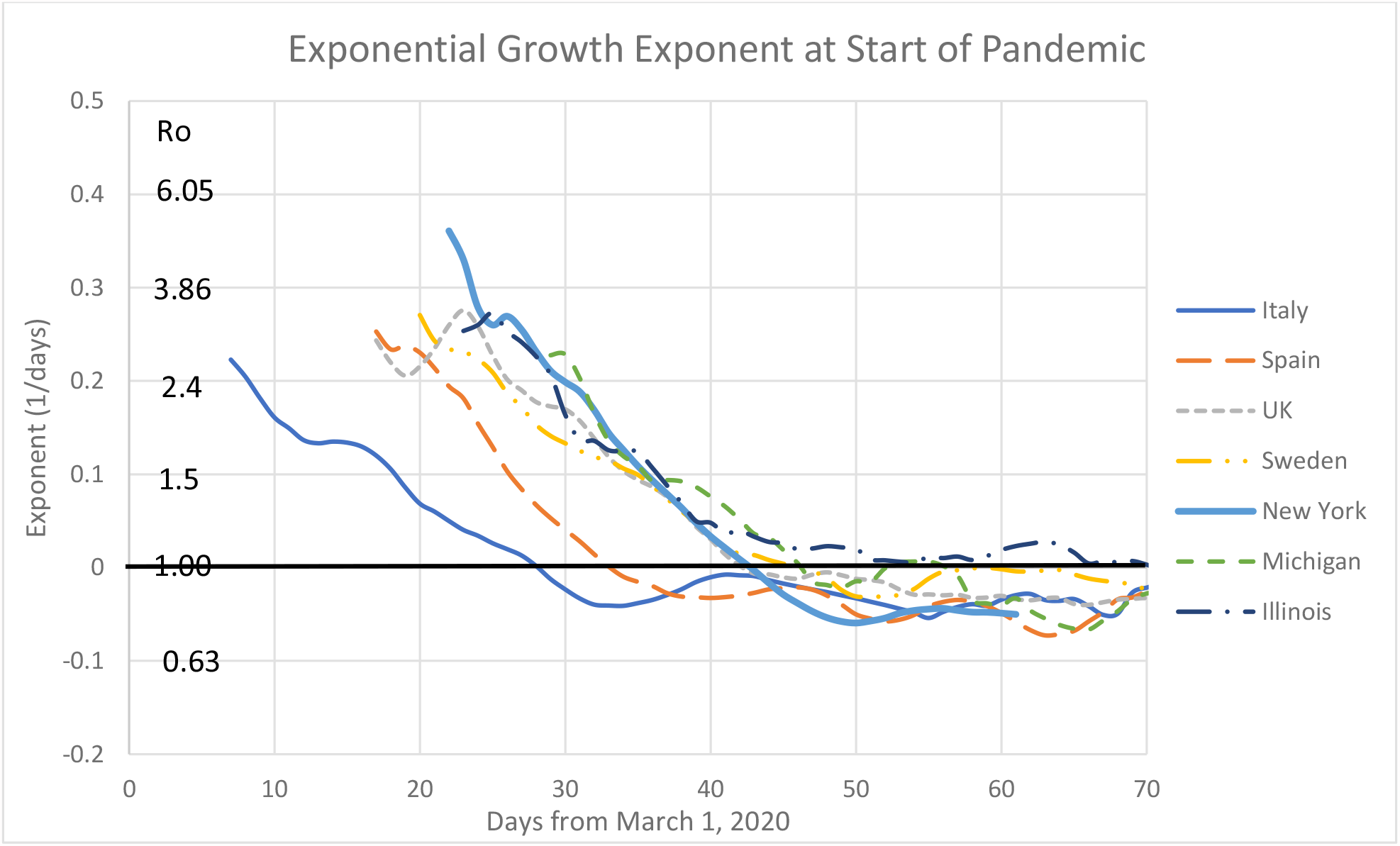
Exponentiating times at pandemic inception (7-day averages)

The numbers along the left side are values of *R*_*0*_ deduced from the approximate relationship *R*_*0*_ ≈ exp (1.5/*κ*), where the exponentiation time is *κT*_*delay*_ and we assume *T*_*delay*_ = 3 days. This relationship was calculated in Appendix A and shown in Figure A2.

The falling curves show how the population in each area reacts to information about the pandemic. The reaction is clear, but there is no evidence that what happened earliest in Italy had any effect on the later response elsewhere. Two weeks later, the initial *R*_*0*_ in New York was even larger than the initial value in Italy.

The rate of public reaction is measured by the slope of the curves in Figure 4. In Italy *R*_*0*_ decreased by a factor of 2 in about two weeks; in New York the response was faster, decreasing by a factor of 2 in about seven days. Yet, the subsequent peak death rate, shown in Figure 2, was a factor of 2.7 higher in New York. The primary cause was the initial value of *R*_*0*_, which was about 2.5 in Italy and 5 in New York. The exponential growth produced by the shorter initial exponentiation time outweighed the faster adjustment.

### The Second Wave

As shown in Figure 2, a second wave of infections and deaths was encountered around the end of the year. The exponentiation times for some of the same nations/states during Dec. 2020 and Jan. 2021 are shown in Figure 5.

**Figure 5.**
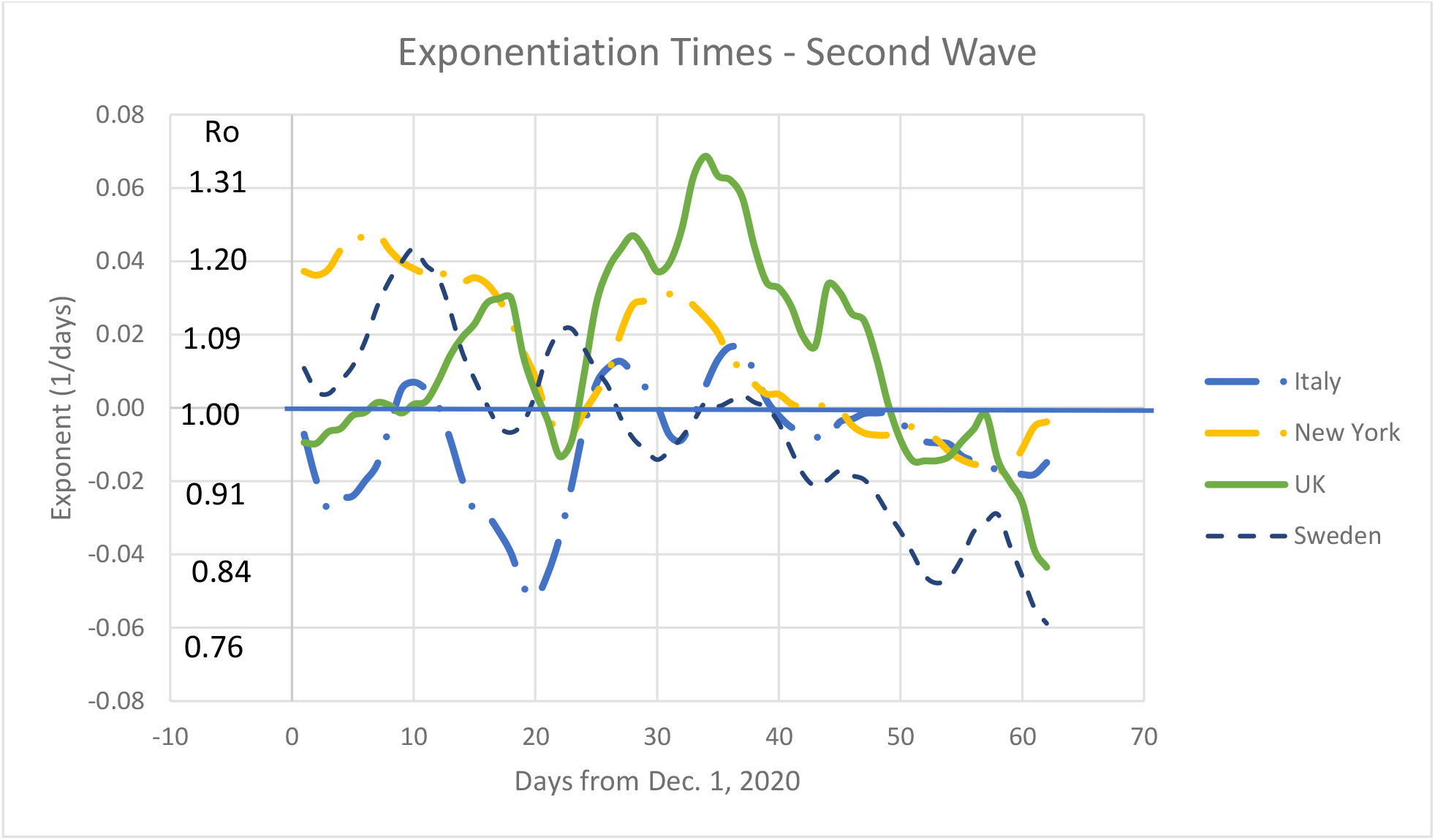
Reciprocal exponentiation times - Dec. 1, 2020, to Jan. 31, 2021 (7-day averages)

The *R*_*0*_ at the pandemic onset shown in Figure 4 decreased from their initial high values in one to two weeks. The *R0* values in the second wave are mostly within 25% of unity and oscillate with periods of one to two weeks. These behaviors are characteristic of a control system in which the sensory feedback signal is delayed by a week or so. The section on Analysis of Community Response in Appendix A modeled such a system by assuming that the community’s average *R*_*0*_ responded to information about tests, hospitalizations and deaths provided by government and the media. That produced oscillations with a much longer period. Oscillations in Fig. 5 must be produced by shorter-term stimuli, e.g., effect of weekends.

It should be noted that *R*_*0*_ remained above 0.75 during the December/January second wave, even when the absolute death rate was still above 5 dpM/day. In all the states and nations reviewed for this analysis, only in Virginia in early March 2021 did *R*_*0*_ decrease to 0.5. Apparently, the community relaxes its social restraints on receiving reports of a decrease in daily positive tests, hospitalizations or deaths, even when the absolute death rate remains at alarming levels.

### States and Nations Reaching an apparent Limit

In June 2020 the worst appeared to be over. Figure 6 shows the accumulated death toll in nations and states where it appeared to have reached an asymptote at about 700 dpM, and for New York State, where it reached about 1250 dpM. The daily death rate had subsided from a peak of 37 dpM/day in New York to less than 2 dpM/day in these areas.

**Figure 6.**
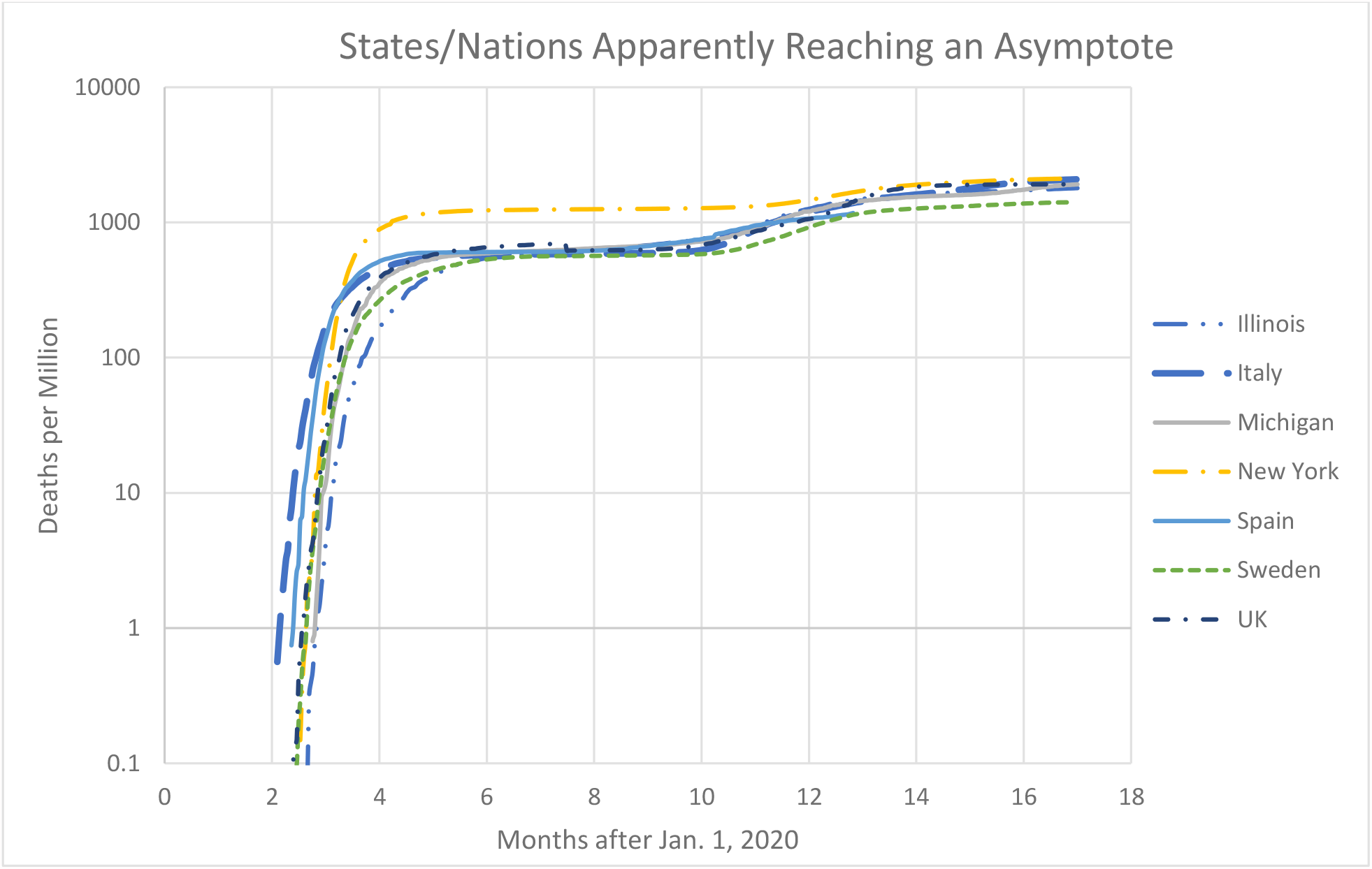
Accumulated dpM for countries and states reaching an apparent asymptote in summer 2020.

This behavior mimics the expected effect of herd immunity illustrated in Appendix A, Figure A1. The credibility of herd immunity being achieved with deaths of about 0.1% of the population depends on *N*_*rec*_, the number of people who survive the infection for each person who dies. Publicized data provided the cumulative number of people with positive test^†^ results for the infection, usually listed as “cases.”

However, the number of confirmed “cases” depends on the number of tests administered and the criteria used for testing, both of which have clearly varied over the course of the pandemic. During the summer of 2020 the ratio of reported “cases” to deaths varied among states from 37 in New Jersey to 196 in Alaska. It is not credible that the quality of medical care varies that much between states, so the difference between these ratios probably reflects differences in local testing policies and the availability of testing resources. Therefore, these values represent lower limits to *N*_*rec*_.

A better estimate of *N*_*rec*_ can be deduced from seroprevalence surveys, which consist of tests for COVID-19 antivirus administered to randomly selected persons in a designated area. Each test determines whether a person is or has been infected. A search of the Centers for Disease Control and Prevention website for seroprevalence data described three types of surveys: large-scale geographic, community-level, and special population [1]. Only the community-level surveys use random sampling to choose the subjects, so only they can produce a reliable estimate of *f*, the total fraction of the population that is recovered and immune in the chosen area. *f* can then be compared with the fraction of the population that died, *d*, for the same area to estimate *N*_*rec*_. Because *d(t)* reflects *f* at an earlier time (approximately a month earlier), the comparison is best made after an asymptote has been reached.

Although many serological surveys must have been performed, very few were reported. Data were available for ten areas tested prior to May 1. 6.9% of the tests proved positive in the New York City area surveyed between March 23 and April 1 [1], although the number of confirmed “cases” at the same time was only 53,803. Thus, the total number of inhabitants estimated to be immune in New York City on April 1 was 12 times the number of confirmed “cases” reported for the surveyed area at the same time. This proves that most victims of COVID-19 in New York City recovered without being counted as “cases.”

To estimate *N*_*rec*_, we need to know the number of deaths that resulted from these infections. However, the death rate in New York was increasing rapidly on April 1, 2020, and some of those who tested positive would die during the subsequent interval between infection and death, *T*_*die*_. We estimate *T*_*die*_ to be approximately 25 days based on fragmentary data, such as the first death attributed to the Sturgis motorcycle rally [2] held in South Dakota from August 7 to 16. It is a reasonable number since many patients had been admitted to a hospital for weeks before recovering or expiring.

The report for the entire New York City area [3] cites 70,637 confirmed “cases” and 2632 deaths on April 1 and 12,781 deaths on April 22. Assuming the measured ratio of 12 between “cases” and infections applies to all of New York City, between 2,632 and 12,781 deaths were caused by the 848,000 infections deduced for April 1, yielding a probability of death after infection between 0.31% and 1.5%.

A more accurate estimate was provided later by Dr. Anthony Fauci during his congressional testimony on September 23, 2020 [4]. He quoted a value of 22% for the fractional immunity in New York City at its temporarily asymptotic death level. Combining this value with the asymptotic death fraction of 0.13% (Figure 6), we deduce that N_rec_ = 169, for an average probability of death after infection of 0.6%, which compares with a typical value of 0.13% for the seasonal influenza. Dr. Fauci’s estimate implies that by September 23, 1.8 million of the residents of New York City had acquired immunity, while the number of confirmed “cases” at that time was only 0.24 million for a ratio of 7.5. Clearly, most people who acquired immunity never displayed symptoms and were never tested for the virus. Whether they were infectious at any time was not known.

This presented a dilemma: although the shape of the accumulated death curves in Figure 6 strongly suggested herd immunity, the 22% infected fraction reported by Dr. Fauci was clearly insufficient to produce it, as illustrated by the calculations in Appendix A. All the curves in Figure 6 exhibit the same behavior, yet none appear to have reached a high enough value of *f* to achieve herd immunity.

To resolve this inconsistency, we postulated that the populations in these areas did not respond homogeneously to the pandemic. Rather, they could be approximated as a hybrid population composed of two sub-populations: those that comply with social restrictions (compliers) and those that deny that there is a problem and defy social restrictions (deniers). That most people were compliers was obvious from viewing the streets during the height of the pandemic. Evidence for the existence of deniers appeared each time the formal restrictions were relaxed: some groups participated in dense gatherings at bars, church celebrations, political rallies, motorcycle rallies, etc. Since these people ignored the risks of contracting the virus by defying guidance when the restrictions were relaxed, it was reasonable to assume that their private behavior prior to the lifting of formal restrictions also ignored the risk: that is, they gathered in dense groups in private quarters instead of public locations. The example set by the President and the associated political polarization also encouraged deniers to ignore restrictions, with many even refusing to wear face masks to protect those they encountered.

The model developed in the Hybrid Population section of Appendix A demonstrates that in a hybrid population with 80% compliers and 20% deniers an asymptotic immunity of 22% could produce a dramatic decrease in the infection rate as the denier sub-population achieves herd immunity. It could even extinguish the pandemic altogether if all remaining susceptible people controlled their behavior so that *R*_0_ (1- *f*) < 1.0. Nevertheless, while a few active infections are sustained, relaxation of restrictions can relight the fire.

### States with Various Reponses^‡^

Figure 7 presents the accumulated COVID-19 deaths per million inhabitants in states with various pandemic growth histories. The earliest onsets were in New York and Louisiana. New York has the largest influx of traffic from Europe. New Orleans, LA hosted the Mardi Gras celebration on Feb. 25, 2020. The death rate in Louisiana reached 10 dpM/day on April 1, 2020, after a delay about equal to the typical time between COVID-19 infection and death. The latest onset was in Hawaii, the state farthest removed from Europe. It has the most interchange with Asian nations, but travel from China had been curtailed by the President on Jan. 31, 2020.

**Figure 7.**
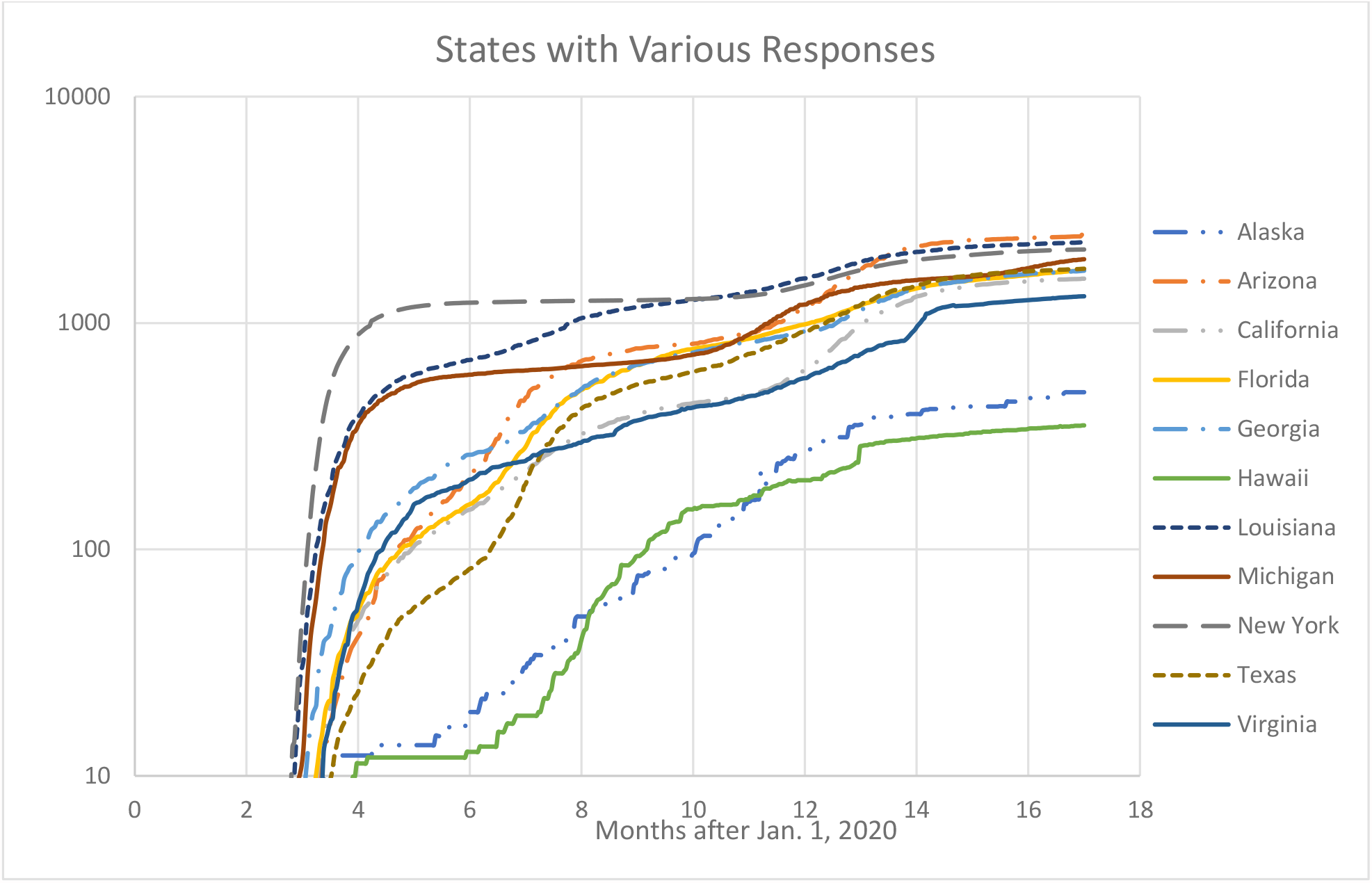
Accumulated COVID-19 dpM for selected U.S. states with different pandemic growth histories.

New York suffered the most from the first wave of the pandemic despite the “New York State on Pause” order issued by Gov. Cuomo on March 22, 2020, to shut down non-essential businesses. By this time New York already had 58 deaths. The President had issued an order on March 12 to restrict travel from UK and Ireland, but not Italy, and it was not applied to U.S. citizens.

New York’s experience, which taxed its medical care and mortuary systems to their limits, served as a warning to other states. Its initial exponentiation rate of 0.36 day^-1^, its eventual peak death rate of 38 dpM/day and its first-wave asymptotic deaths at about 1200 dpM were all among the highest in the world.

A slower initial growth rate enabled Louisiana to slow down the pandemic initiated by the Mardi Gras celebration near the 600 dpM asymptote, but persistent death rates between 2 dpM/day and 10 dpM/day accumulated an eventual total of 2270 dpM, which surpassed even that of New York.

The Michigan curve mimics New York, but with smaller first-wave asymptote at 600 dpM and eventual deaths at 1900 dpM.

Georgia is an outstanding example of politics overriding good sense. It was among the leading U.S. states in deaths from the outset, but officials still fought over restrictions; there was even a legal battle between the Governor of Georgia and the Mayor of Atlanta over face-mask requirements. Defenders could argue that their eventual toll of 1800 dpM is no worse than many other states.

California provides an example of early control followed by relaxation after many months of sacrifice. Gov. Newsom issued a “Stay at Home” order on March 19 to restrict the size of the gatherings. The President’s cutting off travel from China with a two-week quarantine for citizen returnees helped prevent the rapid onset experienced by New York. However, the number of deaths continued to increase gradually until the Thanksgiving, Christmas and New Year celebrations caused the accumulated death toll to reach 1570 dpM. It appears that the principal effect of the early restrictions was to postpone the eventual deaths. The resulting impact on the economy remains to be evaluated.

Florida, Texas and Arizona have similar histories. Initially, they were favored with relative low rates of infection. However, overconfidence and political controversy produced an ongoing high death rate during the summer of 2020 and the second wave at year’s end. The eventual accumulated deaths in Arizona at 2420 dpM exceeded even New York’s.

After an initial surge, Virginia has managed to maintain a relatively low death rate except for a surge at the end of February 2021. Its final death toll was 1315 dpM, 55% of Arizona’s value.

Hawaii and Alaska demonstrated how their natural separation could be used to protect their population from the pandemic. They limited the eventual deaths to 355 dpM and 495 dpM, respectively. Both insisted that entrants maintain self-quarantine for two weeks after arrival in the state, while strongly recommending that people wear face marks and maintain social separation.

### Selected Nations

Figure 8 presents the accumulated COVID-19 death fraction for various nations.

**Figure 8.**
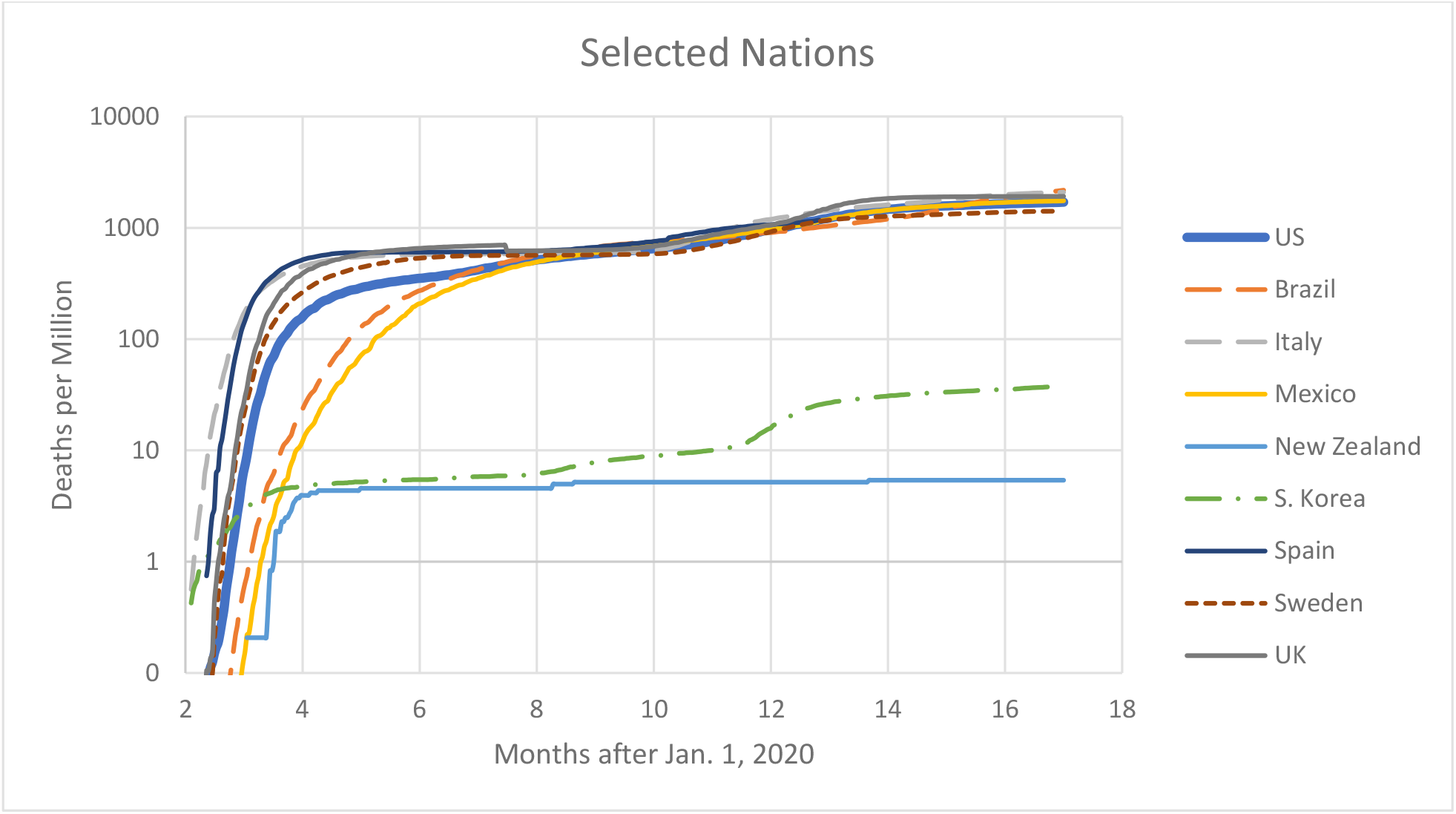
Accumulated COVID-19 dpM for various nations.

S. Korea imported the COVID-19 virus directly from China, but it clearly applied the most effective response, as discussed above. New Zealand banned travel from China pre-emptively in early February 2020. The first confirmed case arrived in New Zealand from Iran on Feb. 26; the second case arrived on March 4 from Italy. The borders were closed to non-residents on March 19 and a general lockdown imposed on March 25. As a result, the death rate subsided to a negligible level by the end of April and domestic restrictions were gradually relaxed in the fall 2020.

Italy was the first western-hemisphere nation to be infected, but most others followed a similar course with similar results. The two waves discussed above are evident in all nations, while the onset was generally delayed by an amount determined by the intensity of travel from those nations with prior pandemics. Transmission to all northern nations in the western hemisphere occurred within a month. Transmission to Brazil and Mexico was delayed by about two months, but their deaths eventually caught up. Brazil accumulated the highest death toll of 2190 dpM on May 31, 2021.

Sweden, which applied the least formal restrictions on public behavior, produced the lowest eventual death toll in these nations, i.e., 1410 dpM. The government’s approach appeared to be to inform and advise the public without applying legal restraints.

### Current Status

Effective vaccines against COVID-19 were introduced in Dec. 2020, but their deployment was too late to prevent the second wave of infections and deaths. Fortunately, they are available to prevent a third wave, but only if a sufficiently large fraction of the population is vaccinated.

According to Fig. 1, total deaths in New York reached almost 1900 dpM by the end of May 2021. Using the inferred infections/deaths of 169, about 32% of the New York population has recovered from the infection. Meanwhile, about 45% were vaccinated. Thus, about 63% of the New York state community are presumed to be immune and the effective *R*_*0*_ is reduced to 37% of its normal-behavior value. As shown in Figure 4, the initial value of *R*_*0*_ in New York was about 5. Therefore, a return to normal behavior would result in *R*_*0*_ ≈ 1.9, or a pandemic increasing with an exponentiation time of about 6 days. Meanwhile, *R*_*0*_ has increased by an unknown factor by the introduction of the more infectious delta variant. Similar analysis of other states reveals a common conclusion: ongoing restraint in interacting with others is essential.

If *R*_*0*_ = 5.0 is typical of near-normal behavior, more than 80% of the population must be immune by a combination of recovery and vaccination before it is safe to return to normal life. The political climate in the U.S. appears to be limiting vaccinations to about 50% of the population; therefore, ongoing infection waves are likely until more than 60% of the unvaccinated part of the population has been infected and recovered. Since the ongoing pandemic has enabled more virulent strains of COVID-19 virus to develop, the current normal-behavior *R*_*0*_ nay be even larger and refusing vaccination may be even more dangerous. Allowing the pandemic to continue anywhere in the world also risks allowing a mutation to develop that resists the vaccines and the antivirus in persons recovered from previous infection, which could reignite an even worse disaster.

## Summary

The COVID-19 infection is transmitted primarily by human-to-human encounters, especially inhalation of air exhaled by an infected person. The infectious agent is carried along with the air on miniscule particles that are too small to be influenced by gravity. Face-to-face conversations are probably the most effective means to transmit the infection.

As demonstrated in Appendix A, the development of a pandemic is determined by the infection multiplier, *R*_*0*_. If *R*_*0*_ < 1.0, the rate of infection, hospitalization and death decreases; if *R*_*0*_ > 1.0 they increase at an exponentially increasing rate. *R*_*0*_ is under the direct control of the members of a community: it is determined by the frequency of person-to-person encounters and the probability of passing on an infection (e.g., proximity and masking) during an encounter. It is also influenced by the virus’ virulence, especially the amount of virus that needs to be passed to a recipient to produce an infection. Halving that amount by a mutation in the virus automatically doubles *R*_*0*_ for a fixed encounter situation.

Analysis of the data on COVID-19 deaths during 2020 and 2021 demonstrates:

> The observed behavior of the COVID-19 pandemic is consistent with results of SEIR model calculations, recognizing that *R*_*0*_ is adjusted by the population in response to data and guidance.
>
> The initial values of *R*_*0*_ ranged from about 3 to 5.
>
> The populations adjusted their behavior so that *R*_*0*_ decreased by a factor of 2 in one to two weeks until it was slightly below 1.0. Thereafter, it tended to oscillate between values slightly below and above 1.0 with periods of 1 to 2 weeks.
>
> Information about the pandemic from the nations affected earlier (e.g., Italy) had negligible effects on the initial response in areas infected later (e.g., UK and New York).^§^
>
> On average, each death was associated with an estimated average of 169 infections, but the reported number of “cases” accounted for less than half of these infections in almost all nations and U.S. states except Alaska.
>
> The apparent long-term periodicity (about 6 months) in the death rates are likely to be caused by delays in the “control loop”, e.g., the population’s reaction to pandemic information. The six-month period is consistent with model calculations using reasonable assumptions about the delay in providing information to the public and in the population’s response to that information.
>
> The short-term periodicity (1 – 2 weeks) must be due to some other factor, perhaps the influence of weekend activities.

## Discussion

Consider the day the first person in New York died of COVID-19, presumably because he/she was infected an estimated 25 days earlier. An average of 168 other people were infected at the same time. By the time we were able to measure the COVID-19 exponentiation rate it was 0.36 day^-1^ (i.e., the cumulative deaths were e-folding every 2.8 days). If this earliest measured rate represents the rate during the previous 25 days, for each of these 169 persons infected on day 1 another 8100 persons had been infected by day 25. Therefore, on the day that the first COVID-19 death occurred in New York, 1.4 million people had already been infected in New York (about 7% of the population) and 8100 of them were fated to die. This could not have been prevented unless the population responded to warnings: e.g., predictions based on Italy’s experience three weeks earlier.

The development of a pandemic such as COVID-19 is determined by the population’s behavior, i.e., the rate at which people encounter each other and how closely they interact. The infection multiplier, *R*_*0*_, can be halved simply by halving the encounter frequency, or maintaining enough separation to halve the probability of transmitting an infection. It can be doubled by a virus mutation that doubles the probability of infection in each encounter.

A particularly dangerous aspect of COVID-19 is that most are infected, and presumably infectious, while not displaying any symptoms, i.e., most infections are transmitted unwittingly! Many who believe they have been responsible citizens and avoided infection may, nevertheless, be infectious and potentially deadly to more vulnerable citizens.

The analysis presented above demonstrates a typical behavior of citizens in democracies:

> They adjust their behavior only to a minor degree until the threat becomes apparent in their interaction area. Some deliberately defy instructions as a demonstration of their “freedom”.
>
> Eventually most respond to convincing evidence of the threat, but that response is delayed enough that the exponential growth continues for a while, producing an overshoot in the infection rate.
>
> They re-adjust their behavior when the threat appears to be abating, but do not persistt until it is suppressed. As a result, there are sufficient infectious people in the population to re-ignite the exponential growth as soon as *R*_*0*_ > 1.0.
>
> The natural result of such behavior is oscillation in the infection rate, i.e., repeated waves of infection and death rates separated by months, depending on the specific reaction times and rates.

The political climate in the US contributed additional reluctance and delay into behavior adjustment. It is incomprehensible to us that political loyalty would persuade people to increase their contribution to *R*_*0*_ by a significant factor by eschewing masks and attending rallies.

Autocracies can prevent such oscillations, as demonstrated in China during the spring of 2020. Wuhan was clearly the source of the COVID-19 virus, and its epidemic initially grew rapidly. Apparently, the government imposed strict discipline, effectively imposing total isolation on buildings in the city and placing a cordon around the city. As a result, the total death toll, even adjusted for suspected manipulation of data by the Chinese government, was far below that reached eventually in the major democracies.

There are alternative approaches to controlling the pandemic, such as that demonstrated by S. Korea and New Zealand. S. Korea received the earliest cases exported from Wuhan into a closely interacting church community. Yet, it managed to limit its accumulated death toll to 37 dpM, a factor of 46 less than the U.S. It had prepared itself with test kits and trained personnel to detect and track all those who might have been infected by the initial cases. It had the determination and authority to isolate all who were infected. This approach can be highly effective, but it must be applied before exponential growth overcomes the available testing and tracking resources.

The U.S. was not prepared. Our only recourse was to slow the importation of the infection by quarantine of arrivals from infected areas. The President suspended entry of non-citizens from China on Jan. 31, 2020, thereby delaying the onset of the pandemic in the U.S. west coast, but entry from Europe was not curtailed until March 2020. The resulting death rate in California during the first wave peaked at less than 2 dpM/day, whereas in New York it exceeded 37 dpM/day.

The states of Hawaii and Alaska made use of their relative isolation to limit deaths by applying moderate restrictions on public behavior and two-week quarantines on arrivals from elsewhere. As a result, they accumulated 355 and 494 dpM, respectively, as compared to 1700 dpM for the U.S. average. For comparison, deaths in the U.S. from the annual influenza season ranged from 70 to 185 dpM during the seven winters prior to 2020.

Anyone who doubts the effectiveness of social distancing should review the data on annual influenza deaths. They were reduced essentially to zero everywhere in the northern and southern hemispheres during the 2020-2021 season, presumably by the social distancing measures taken to control COVID-19.

## Lessons Learned

The important question is, “How should we prepare to meet future challenges similar to COVID-19?”

### Guide Nationally, Manage Locally

The variety of responses to the pandemic in different areas and the differences in response to the first and second waves show that applying remedies (e.g., social and business restrictions) nationally is wasteful and demoralizing. There are vital tasks to be performed for the nation, but managing the pandemic should be more local, preferably by response area (e.g., area with established management in which people interact frequently). The response area could be as large as a state (e.g., Wyoming) or as small as a borough in New York City.

Tasks requiring national leadership include:

> At all times, stockpile equipment and supplies required to control pandemics at their inception.
>
> In preparation, provide training for personnel needed to control pandemics at their inception.
>
> In anticipation, provide warning and general advice to the public. Firmly worded advice to wear masks is appropriate. Masking impacts little discomfort and decreases *R*_*0*_ by a significant factor, which is particularly beneficial at its onset. Otherwise, avoid imposing severe constraints on areas that have not been infected for fear of producing resistance later when they become important.
>
> Monitor closely and publicize the development of infections, hospitalizations and deaths in all areas. Perform and publicize seroprevalence surveys to guide the model calculations.
>
> During the pandemic, provide ongoing guidance and data to local managers.
>
> Provide data, such as models and key parameters to support analysis and predictions, such as the model provided in Appendix A.
>
> Perform research to resolve uncertainties in models, such as the parameters used in the model.
>
> Conduct seroprevalence surveys to monitor the progress of the pandemic and publish the results.
>
> Provide up-to-date data on test results, hospitalizations, deaths, etc.
>
> Produce and distribute important equipment and supplies to the most needful areas, precluding a bidding war between desperate users.
>
> When necessary, invoke the Defense Production Act to accelerate the availability of essential items.

Tasks that are more appropriately performed by the local (response area) management are:

> Inform the local population of the intensity of the threat. Use data but avoid large numbers that have little meaning to most people. Use comparisons, such as deaths per million compared to annual traffic fatalities (about 100 dpM). Offer predictions of the future as it depends on whether public behavior (e.g., *R*_*0*_) is modified. Build public confidence by showing that the prior predictions were correct. Avoid political issues where possible.
>
> Use prediction results to provide guidance.
>
> Issue enforceable orders restricting activities to prevent an increase in *R*_*0*_ above acceptable levels

### Prepare to Respond Promptly

The data demonstrate that there are two distinct types of responses in each response area (e.g., area in which people interact frequently): control the spread at its inception or minimize it after it has escaped initial control. They also show that long-term, large-scale oscillations are the natural result of delay in feedback control, so the feedback (public reaction by adjusting *R*_*0*_) must be anticipatory instead of reactive.

Control at inception requires:

> A plan with personnel trained to execute it.
>
> Capability to control ingress into the response area.
>
> Adequate supply of test kits and personnel trained to track possible infections.
>
> Legal authority to track infections and resources to quarantine infectious persons.

The same techniques can be applied later once the number of infectious persons has been suppressed by social isolation and vaccination to a level consistent with the available resources.

### Manage Pre-emptively

Minimizing the pandemic in a democracy requires the ability to <u>predict</u> the pandemic’s future and to <u>persuade</u> the public to alter its behavior accordingly.

The tools to predict exist; they only require determining the appropriate value of a few parameters and an estimate of the future value of *R*_*0*_. Data from other nations/states with earlier onsets can also serve to warn the public.

Persuading is difficult in a democracy. The U.S. experience in 2020 proves that persuasion by edict is ineffective and that persuasion by political loyalty is dangerous. Scientists try to persuade each other by formulating a consistent logical argument, but its reception requires an educated, logic-receptive audience. Improving U.S. education could go a long way in this direction, but that is a long-term solution.

In the near term, persuasion by experience may be the best available. Certainly, the U.S. suffered severely enough from the 2020-2021 COVID-19 pandemic to get people’s attention. Yet, a large fraction of the population still resists vaccination, in opposition to all quantitative data. Nevertheless, a rational approach by leaders, one that makes verifiable quantitative predictions, should eventually prevail over emotional appeals. The formula is a simple repetitive cycle:

1. The model is used to predict the future effects of the pandemic based on current behavior.
2. The leader informs the public of the consequences of maintaining current behavior and recommends modifications.
3. The public ignores the recommendations.
4. The public suffers the consequences, as predicted.
5. The model is recalculated with updated information.
6. The leader informs the public of the consequences of the new situation and recommends behavior modifications.

Eventually, this process should generate confidence in the model and the leader, while creating enough social pressure on the dissenters to persuade them to comply.

## Limitations

The data chosen for the analysis were for selected states and nations, for which we believed the available published data to be reliable. We avoided nations suspected of manipulating data for political reasons (e.g., Russia, China). We also excluded others for which the results were similar to those of included countries. Iceland, for example, suppressed the pandemic at its inception by methods like those employed in South Korea and New Zealand and by taking advantage of its relative isolation.

## Conclusions

The most important conclusions to be drawn from these data are as follows.

> Most COVID-19 infections are transmitted from people who are not aware of being infectious.
>
> In the U.S. and western Europe all attempts to arrest the pandemic at its inception by behavior restrictions failed. The lowest fatalities were in Sweden, where the government focused on informing the public rather than imposing regulations.
>
> Antivirus survey tests indicate that, on average one person died for each 169 persons infected, giving a probability of death of 0.6%. Less than half of those with positive antivirus tests had been counted as “cases”.
>
> The apparent asymptote in deaths in the summer 2020 was the result of attaining herd immunity in the about 20% of the population that defied restrictions, or it was the first phase of long-term oscillations.
>
> The second wave experienced at the end of 2020 was the manifestation of long-term oscillations caused by the population reacting to information on the pandemic, information that is significantly delayed from the infecting events.
>
> Unless a sufficient fraction of the population is vaccinated and a virus variant does not defeat the vaccine, we predict another serious infection wave in September 2021.

The restrictions imposed in some states, but apparently not obeyed by a minority group, thus had the following effects:

- They protected the medical facilities from overload.
- They prolonged the shutdown of economic activity.
- They enhanced “cabin fever”.
- They failed to arrest the pandemic.

These conclusions lead to the question, “What would have resulted if the U.S. and state governments and acted differently in 2020-2021ã”. Speculations are presented in Appendix B.

According to our model a virus mutation that increases the probability of infection by a factor of 2 will convert the current death rate, which appears to be acceptable to the U.S. population, into an exponential growth with an exponentiation time of about 10 days. Since it takes about 25 days for the infection rate to appear in the death rate, by the time the renewed growth is apparent the infection rate will already have increased by a factor of about 12.

## Data Availability

The data are all available on the Internet.

## Acknowledgment

The author gratefully acknowledges the enormous contribution of Dr. Neal J. Carron for his very careful review, checking the mathematics, and suggesting clarifying improvements.

## Appendix A PANDEMIC MODEL

### Mathematical Model

A model of the COVID-19 pandemic is based on the following assumptions and definitions:

- The infection is spread primarily by an encounter between an infected and an uninfected person.
- The newly infected person becomes infectious after a period *T*_*delay*_, typically 3 days [5].
- The infected person is infectious for a period *T*inf before he/she is removed from the interacting general population by quarantine or hospitalization. We assume that the person is removed within one day after the onset of symptoms. Symptoms appear approximately 5 days after infection, so *Tinf* typically lasts for 3 days. [5]
- The fraction of the population that has acquired the infection at time *t* is *f(t)*.
- The rate at which the average person encounters others is *µ. µ* is 0 if an individual never interacts closely with another person; it may be several per day or hundreds per week if he/she is engaging in normal life.
- The probability that an encounter will transfer the infection from an infectious person to an uninfected person is *P*.

*µT*_inf_ is the number of persons encountered by an infectious person, and *R*_*0*_ *= µPT*_*inf*_, the infection multiplier, is the average number of people that each infectious person infects during his/her infectious period.

Therefore, at the beginning of the pandemic, when *f* is near zero, *Ro* others will be infected by each infected person. It is obvious that the pandemic will expand or shrink according to whether *R*_*o*_ is greater or less than 1.0: if *Ro* is greater than 1.0, the rate of increase of infections, d*f*/dt, will grow until *f* becomes large enough to reduce the effective multiplication rate below 1.0 (because encounters with previously infected persons are assumed not to create new infections).

Thus, *R*_*o*_ *= µPT*_inf_ is a critical parameter, the infection multiplier. It consists of three independent factors, each of which is under people’s control, in principle.

Consequently, before developing any equations, recommended ameliorative actions are obvious:

- Decrease *T*_inf_: isolate infected people as soon as possible by testing and quarantine or hospitalization.
- Decrease *µ*: practice social distancing.
- Decrease *P*: wear masks and maintain separation.

The factors that make COVID-19 particularly dangerous are the following:

- The virus is highly contagious, so that *P* is large unless our behavior is modified.
- There is a period of two or three days after *T*_*delay*_ during which an infected person is infectious but not symptomatic; that is, <u>he/she is not aware of being infectious</u>.

### Development of Controlling Equation

If *N*_0_ is the total initial population, the rate at which the number of infected persons *f*(*t*)*N*_o_ increases is equal to the product of four factors:

> The number of infectious persons, *f(t)N*_*0*_
>
> The rate at which each person encounters another person, *µ*,
>
> The probability that the other person is not already infected or immune (1-*f*),
>
> The probability that an encounter transfers an infection *P*,

First, suppose that once a person is infected, he or she remains infectious forever and that *T*_*delay*_ = 0. Then, the number of infectious persons would be the same as the number infected *fN*_o_ and that number would increase according to

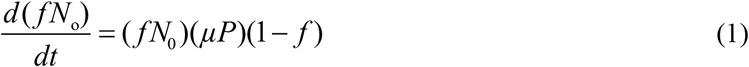

Where *fN*_*0*_ is the number of infected persons, *µP* is the number of encounters per unit time that can transfer the infection, and (1-*f*) is the probability that the other person in the encounter is not immune due to a previous infection.

However, for COVID-19 a person is not infectious until *T*_*delay*_ after his/her infection, and only for a time

*T*_*inf*_ after that. Thus, the fraction of infectious persons is not *f*(*t*) but

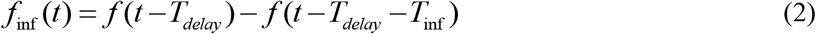

that is, those who were infected between (*t - T*_*delay*_*-T*_*inf*_) and (*t* - *T*_*delay*_), which should replace the first *f* in Eq. 1. Thus, we obtain:

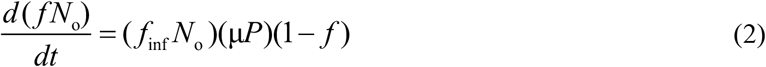

as the controlling equation.

*f*_*inf*_ can be approximated by 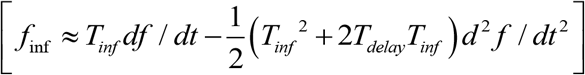 with *f* evaluated at *t*, leading to

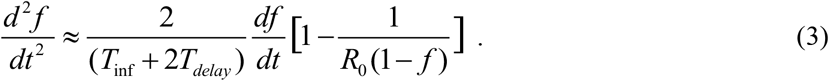

As expected, if the actual infections per infectious person *R*_0_ (1- *f*) falls to 1.0, the second derivative of *f* becomes zero and the rate of new infections *df/dt* becomes constant. Thereafter, *df/dt* decreases toward zero as *f* continues to increase.

In the early stages, when *f* <<1, *f* increases exponentially with an e-folding time *κT*_*del*_ that satisfies the transcendental equation^**^

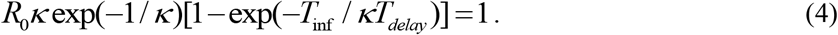

Because we do not know how to solve Eq. (2) or (3), we performed numerical calculations using a spread sheet^††^. Typical values reported by experts are that symptoms appear approximately 5 d after infection and that the person is infectious approximately 2 d prior to presenting symptoms [5]. If we assume that a prudent person will enter quarantine within 1 d after developing symptoms, then *T*_*inf*_ *= 3* d and *T*_*delay*_ *=* 3 d.

### Numerical Solutions

Figure A1 presents the evolution of *f* in time units of *T*_delay_ for different values of *R*_*0*_ and *T*_*inf*_ *= T*_*delay*_. The time scale was determined by *T*_*delay*_. If *R*_*0*_ = 1.0, the infected fraction^‡‡^ remains nearly constant and eventually decreases very slowly.

**Figure A1.**
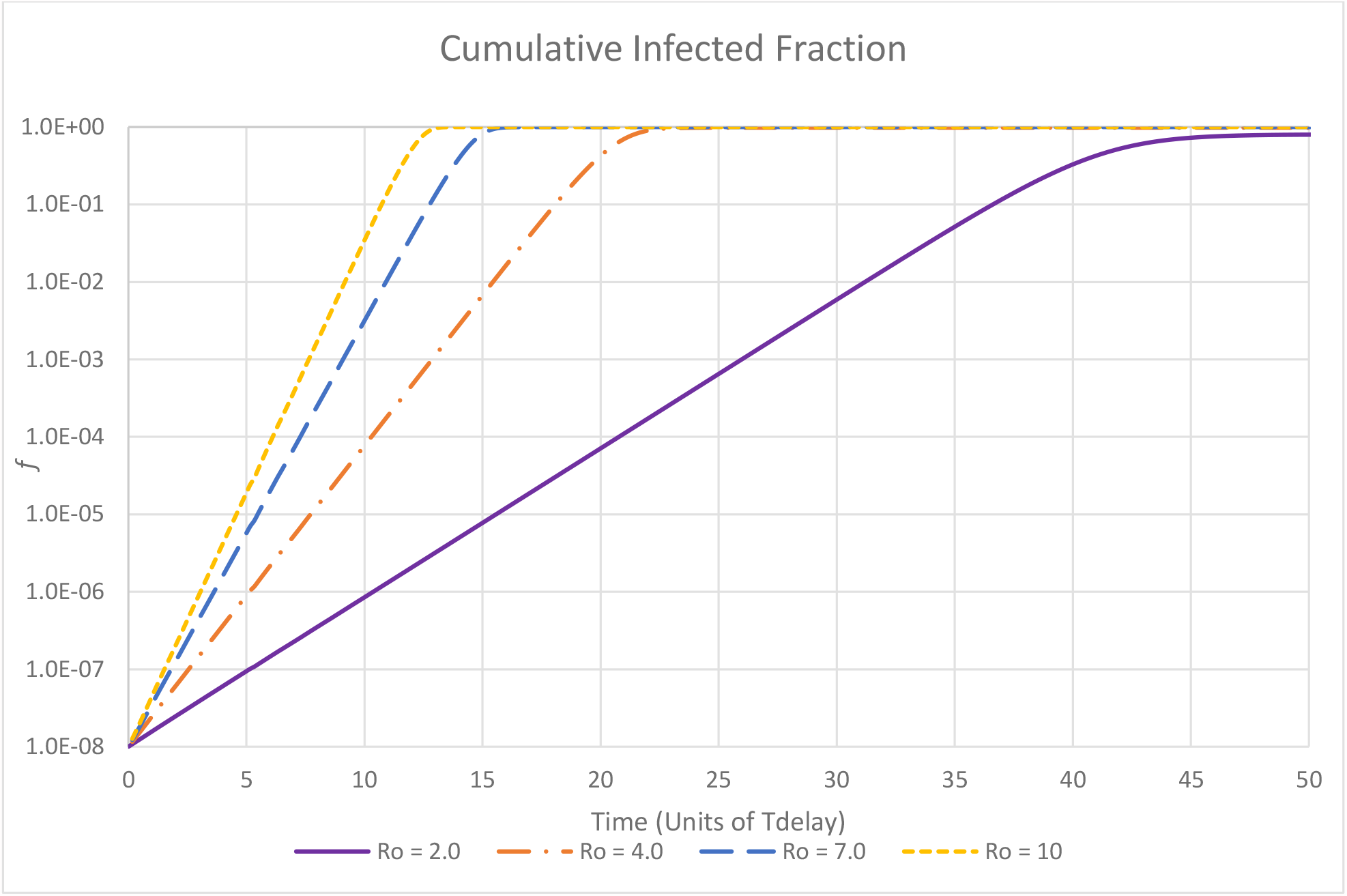
Development of cumulative infected fraction.

Starting from the seed fraction, the infected fraction increases approximately exponentially with time, 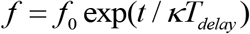 until *f* approaches unity, that is, until a significant fraction of the population was infected and recovered or died, and is no longer susceptible to infection. The exponentiation time, *κT*_*delay*_ decreases with increasing *R*_*0*_. The most useful form of this relationship is its inverse, the dependence of 1/κ, the reciprocal of the exponentiation time measured in units of *T*_*delay*_, as shown in Figure A2

**Figure A2.**
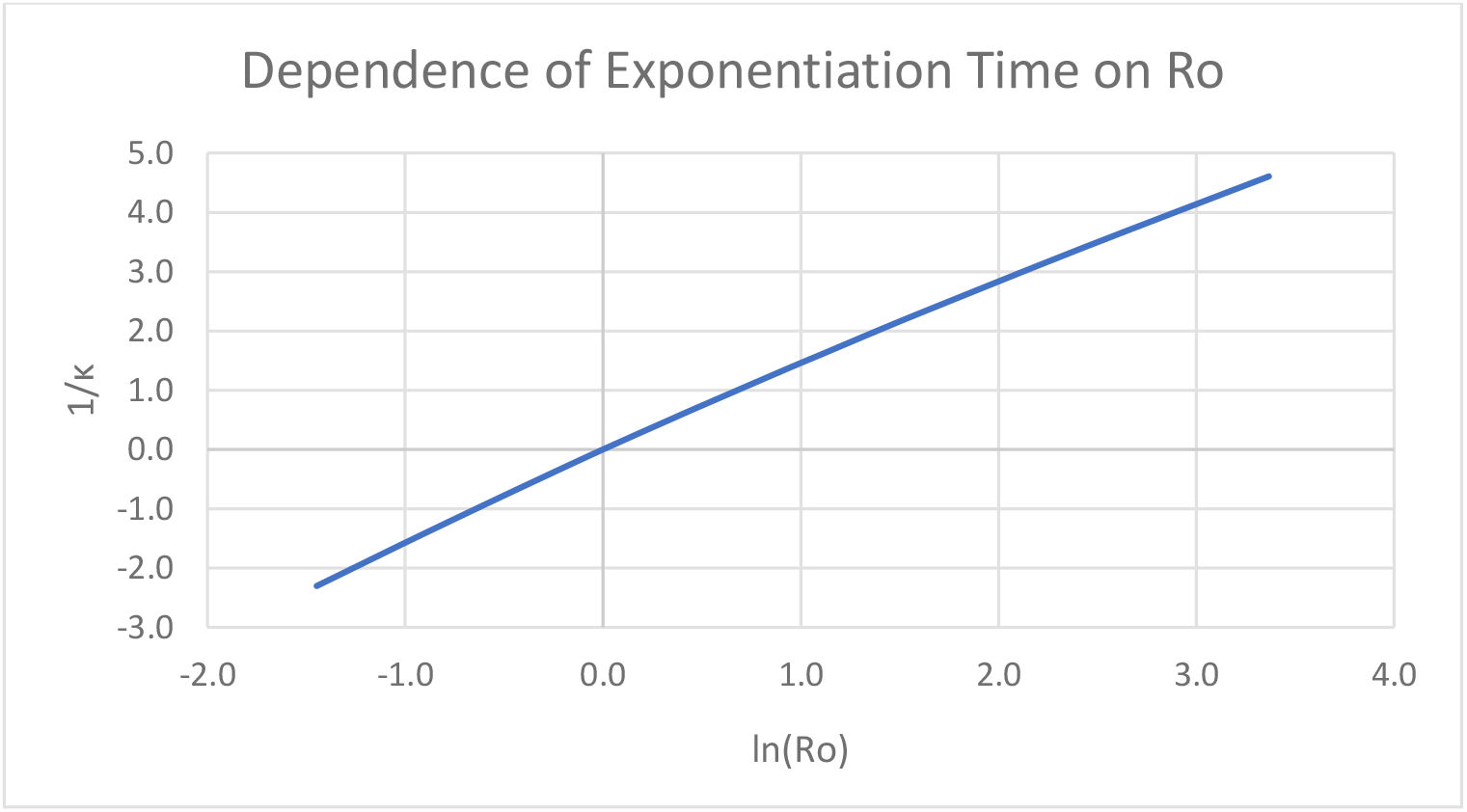
Dependence of reciprocal of the exponentiation time on *R_*0*_*.

The logarithm of *R*_*0*_ is almost - not exactly - proportional to the reciprocal of the exponentiation time, i.e., 1/ *k* ≈ *ln*(*R*_0_) /1.5.

If *R*_*0*_ remains constant, the reciprocal of the exponentiation time, 1/κ, decreases as *f* approaches unity, as shown in Figure A3.

**Figure A3.**
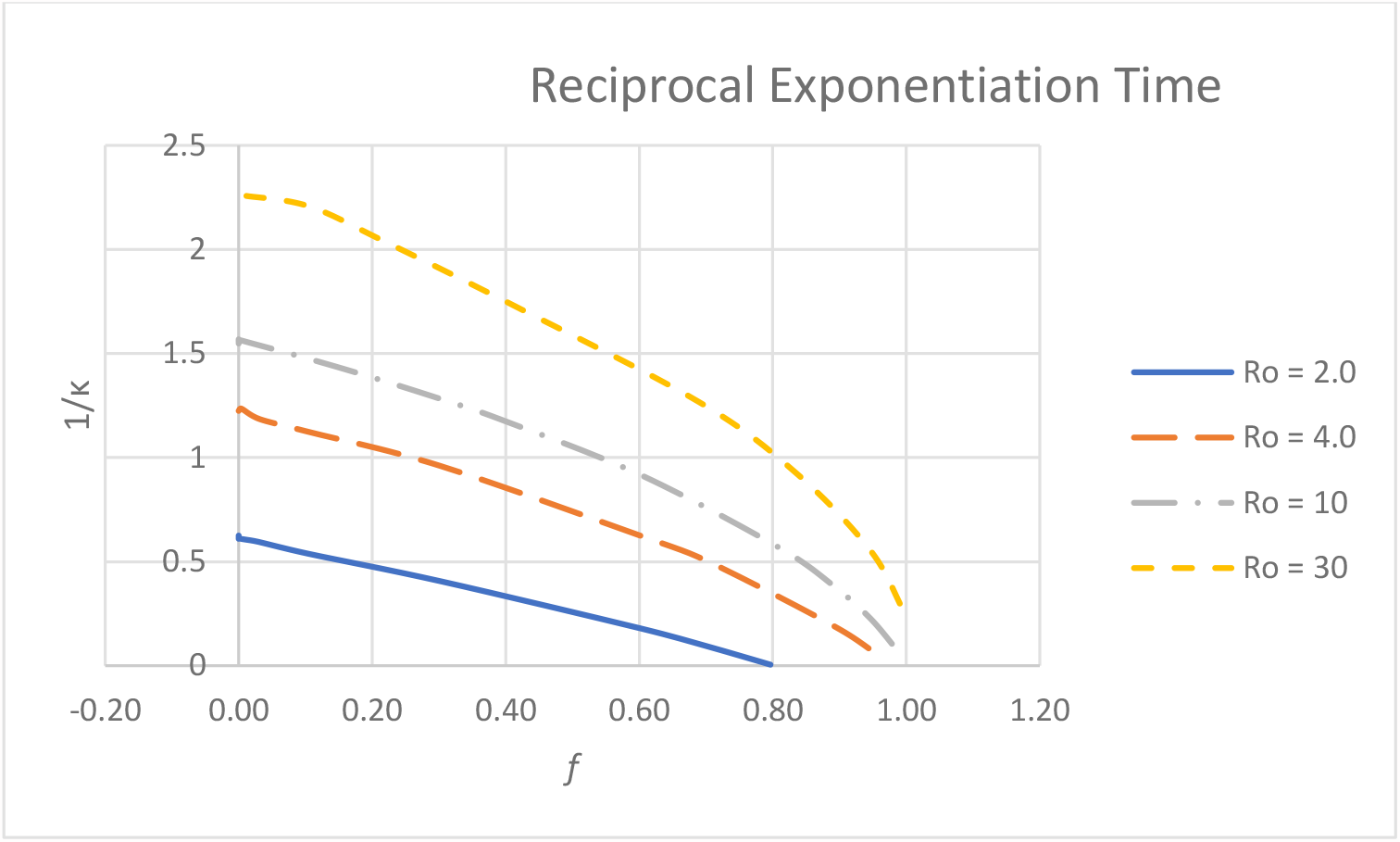
Dependence of reciprocal exponentiation time on f.

Actually, people change their behavior during the course of a pandemic, so high initial values of *R*_*0*_ are usually substantially reduced before half the population has been infected. Thus, the histories of actual reciprocal exponentiation times will fall more steeply than shown in Figure A3 as people respond to alarming data.

The increment in *f* during each unit of time is proportional to the rate of infection, but it is proportional to the rate of deaths at a later time, *t + T*_*die*_. It is shown as the increment during each T_delay_ period in Figure A4. The number of daily deaths can be derived from these curves by multiplying the ordinate by the population and the fraction of the infections that result in death and dividing the result by *T*_*delay*_. It is remarkable that for T_delay_ = 3 d, more than 3% of the population can be infected per day at the peak. This is an implication of exponential growth. In practice, people will become frightened and moderate their behavior, decreasing *R*_*0*_ before it reaches this peak.

These curves are approximately exponential, upward when *f* <<1 and downward when (1-*f*)<<1. The peak rates were reached, that is, herd immunity overcame the effect of *R*_*0*_, when the infected fraction values reached values ranging from 0.43 for *R*_*0*_ = 2.0 to 0.70 for *R*_*0*_ = 10.

**Figure A4.**
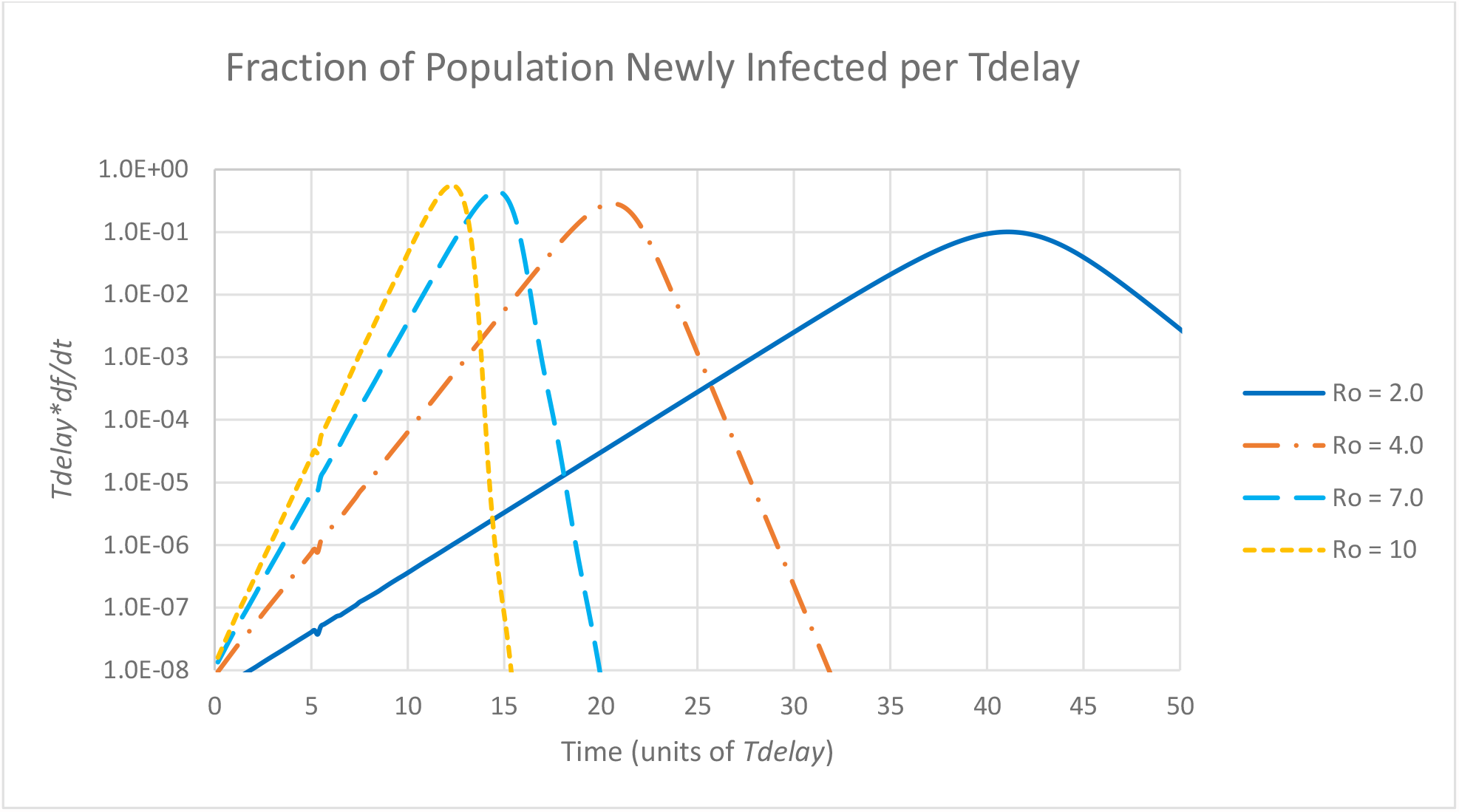
Fraction of population infected during each *Tdelay* interval.

### Hybrid Population

The foregoing calculations show that the rate of pandemic growth is determined by the value of *R*_*0*_, which is the product of three factors: the frequency of interactions between people, the efficiency with which the infection is transferred, and the average time during which an infected person is infectious.

Calculation result presented so far assumed homogeneous populations, that is, they assume that everyone follows similar isolation guidelines. However, experience in the U.S. suggests otherwise. We can model this situation by postulating two sub-populations:

> Compliers: those that take the pandemic seriously and comply with social restriction guidelines.
>
> Deniers: those that deny the seriousness of the pandemic and defy restrictions.

To investigate this situation, we need to divide equation (3) into separate equations for each sub-population. Let α_1_ and α_2_ be the fractions of the total population, *N*_*0*_, represented by the two groups (compliers and deniers), *f*_*1*_*(t)* and *f*_*2*_*(t)* their cumulative infected fractions and *f*_*1inf*_ and *f*_*2inf*_ their respective infectious fractions. *P* is the probability of transferring the infection from an infectious to a non-infected person during an interaction. (We assume the same value for both populations, even though deniers also tend to eschew face masks). *μ*_11_, *μ*_12_, *μ*_22_ are the rates at which members of each population would interact with others if all others were of the same group. Thus, the rate at which a member of group 1 interacts with members of group 2 is *α*2 *μ*_12_.

Equation (3) then becomes:

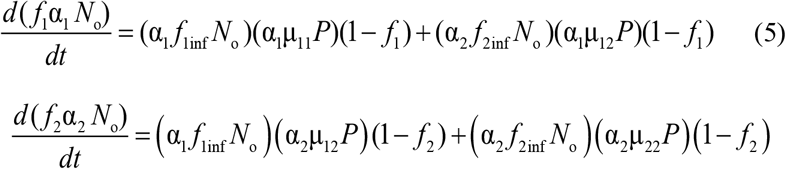

The fraction of the total population that is infected is:

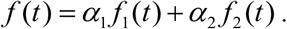

The effects of the different behaviors by the two groups are contained in the µ values.

We investigated the effect of a particular population composition: the compliers, who comprise most of the population, maintain *R*_*0*_ = 1.0, whereas the deniers continue nearly normal interactions, but only with others in their own group. Their value of *R*_*0*_ is adjusted so that the average of the total population is maintained at *R*_*0*_ = 2.0. The complier group on its own would maintain the infection at a constant level since 1.0 person is infected by each infectious person. The results are displayed in Figure A5.

**Figure A5.**
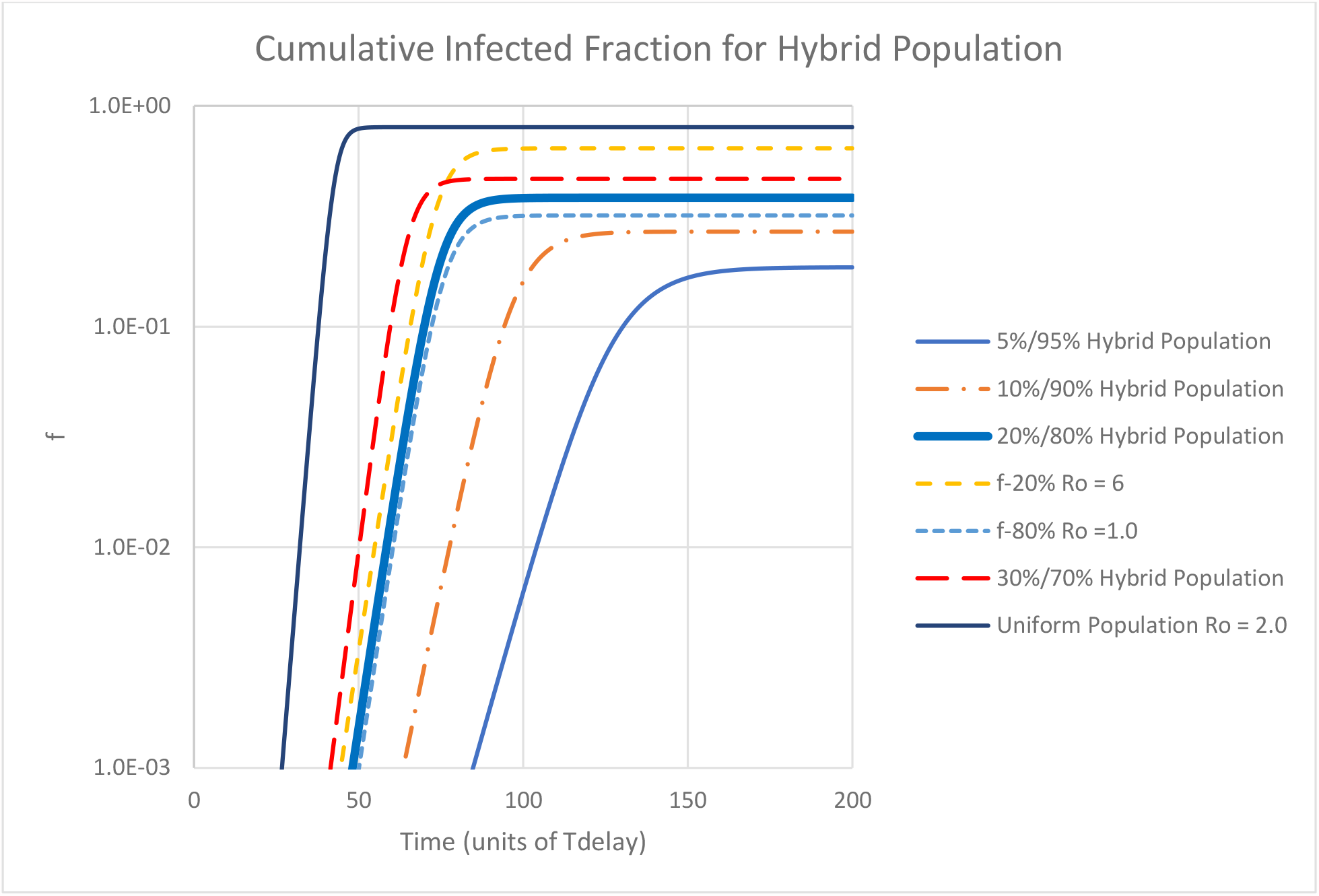
Effect of deniers on population infection with a population average *R*_*0*_ *=* 2.0.

The heavy solid blue curve represents the fraction of the total population that becomes infected if 20% of the population maintains *R*_*0*_ *=6*.*0*, while for 80% of the population *R*_*0*_ *=1*.*0*. The other solid curves represent different denier fractions, each with its *R*_*0*_ adjusted to make the population average *R*_*0*_ *=*2.0. The development of the 80%/20% pandemic is slower than the uniform *R*_*0*_ = 2.0 case, and its asymptotic value of infections is 39% instead of 80% for the uniform population. The short-dashed curve represents the 80% complier population, almost all of whom would not be infected in the absence of deniers. The exponentiation rates for the two populations are identical at 0.21 *T*_*delay*_^*-1*^ since the infection growth is controlled by the denier population. The exponentiation rate is characteristic of a population *R*_*0*_ = 1.36, a value slightly higher than the denier population *R*_*0*_ times its population fraction.

The effect of deniers on the limiting infected fraction and the exponentiation rate, all for populations with an average *R*_*0*_ *=2*.*0*, are shown in Fig. A6.

**Figure A6.**
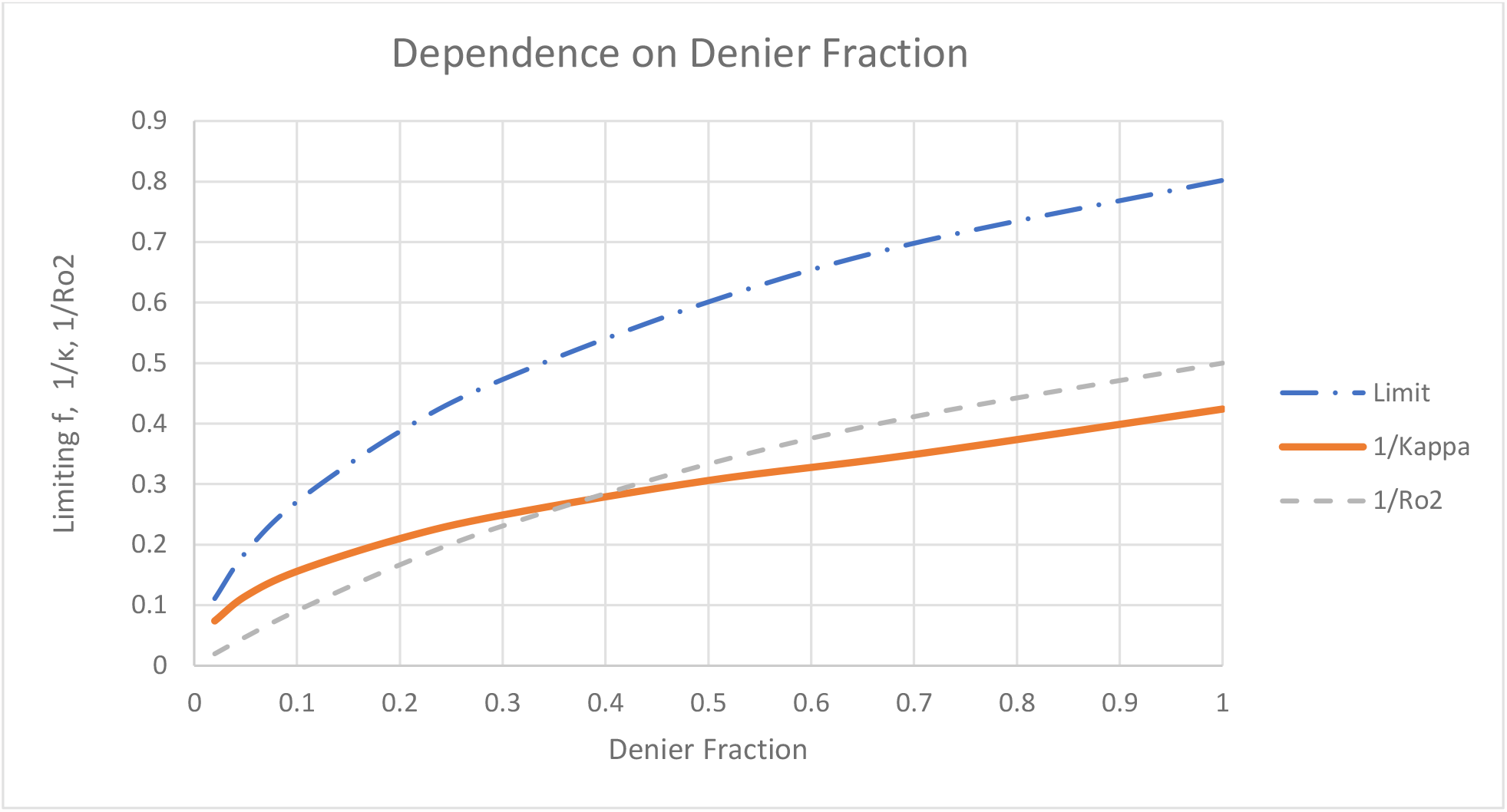
Asymptotic infection fraction, exponentiation rates and 1/*R*_*02*_ required to maintain average *R*_*0*_ = 2.

### Effect of Community Response

The foregoing calculations have assumed that *R*_*0*_ of the population, or parts of the population, is fixed in time, even though it is largely determined by the popular behavior. More realistically, *R*_*0*_ is influenced by the information received by the population, particularly when the data are alarming. The predominant information is provided by official government communications and favorite news sources, which emphasize the most dramatic occurrences. During the COVID-19 pandemic in 2020-2021 the principal data available to the U.S public were the number of positive virus tests, estimates of the number of infections, the reported number of deaths attributed to the virus, and impending crises in hospital staffing, ICU beds and respirators. Data on death rates were the most reliable, but they lagged the infection rate by about a month. Even the hospitalizations occurred a week or two after infection.

A model of community response must deal with this delay. Our choice was to incorporate an adjustment to *R*_*0*_ when the rate of change in *f* evaluated at 5\**T*_*delay*_ previously exceeds a response threshold of .001 *T*_*delay*_*-1*:

> For *T*_*delay*_*df/dt* < .001 *R*_*0*_ remains at its base value, or is increased back toward its base value by a factor of 1.1 per *T*_*delay*_
>
> For *T*_*delay*_*df/dt* > .001 *R*_*0*_ is decreased by a factor of 1.1 per *T*_*delay*_ until it reaches a value of 0.5

The resulting *f* and *T*_*delay*_*df/dt* for *R*_*0*_ *=* 2.0 are compared to the corresponding values without response in Fig. A7.

**Figure A7.**
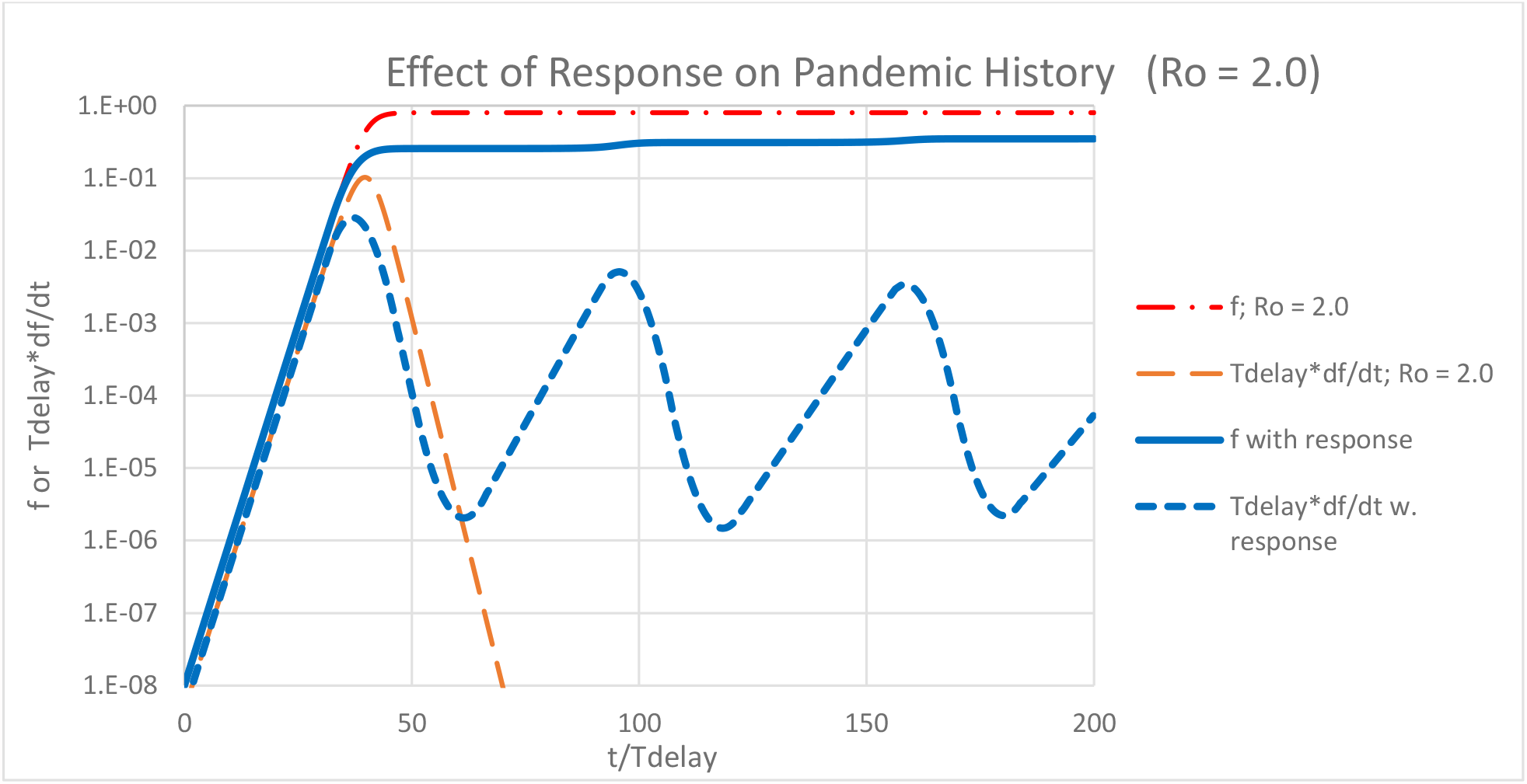
Effect of population response on pandemic history for *R*_*0*_ = 2.0.

This oscillatory behavior is typical of a control loop with delayed feedback: the output overshoots in both the upward and downward directions. The level at which the first plateau in *f* is reached is approximately proportional to the *df/dT*_*delay*_ threshold. The oscillation period is shortened by responding more quickly, e.g., dividing or multiplying *R*_*0*_ by 1.2 rather than 1.1 per *T*_*delay*_.

### Effect of Vaccination

The foregoing calculations demonstrated how the development of the pandemic depended on the value of the effective infection multiplier *R*_0_ (1- *f*). Introducing an effective vaccine in Dec. 2020 added a new factor, *f*_*vacc*_, the fraction of the population that is immunized by vaccination. Since vaccines are administered irrespectively of prior infection, the appropriate means to incorporate vaccination into the model is to replace the factor (1-*f*) in equations (3) and (6) by (1 - *f*) (1-*f*_*vacc*_).

## Appendix B SPECULATIONS on ALTERNATE APPROACHES

### Introduction

Given our model, key parameters and some indications of the population’s response, we can speculate about the outcomes of other approaches to controlling the pandemic. Lack of preparation denied the U.S. the preferred approach, which was demonstrated by S. Korea: contain the pandemic at its inception. Otherwise, we can speculate about the result if the national and state governments had done nothing, imposed restrictions on travelers from Europe as well as China, or if state and local government restrictions had been more or less severe, and/or imposed earlier or later.

We commend the federal government for encouraging the development and deployment of vaccines in 2020. However, it had minimal impact on the pandemic development through May 2021 under any of these alternatives.

We offer a general recommendation for federal action under the threat of a similar pandemic. It should advise, recommend strongly, and/or insist that we wear a mask whenever we expect to come near anyone not in our own family enclave and possibly not vaccinated. If not, we should at least turn our head sideways when we talk with others. Since the primary means by which the infection is spread is by person-to-person contact via transfer of exhaled air into a recipient’s lungs, minimizing the transfer of breath from one to the other reduces *R*_*0*_ by a significant factor. Since there is a delay between infection and onset of symptoms, decreasing *R*_*0*_ is particularly important during the unseen inception phase.

### No Action

The result of taking no action on eventual deaths is clear if we assume, optimistically, that the peak infection rates do not stress the medical system sufficiently to increase the probability of dying. Then the pandemic would have propagated throughout the U.S. until herd immunity was reached when about 80% of the population had become immune. The result would have been 0.80 x 331 million /169 = 1.6 million deaths in the U.S. Any overstress in medical care would increase this number. Despite the record-setting pace of vaccine development and deployment, it would have arrived too late to prevent many of these deaths.

### Control Travelers from Europe and Asia

The President curtailed travel from China on Jan. 31, 2020; in March 2020 he imposed a similar ban on travelers from UK and Ireland. Technically, they applied only to foreign nationals, although immigration authorities encouraged citizens to quarantine themselves for two weeks. Even though the earliest infections were detected in Washington state, the pandemic remained relatively mild in west coast states throughout its first wave. On March 19, 2020, Governor Newsom of California and on March 22 Governor Cuomo of New York issued orders intended to restrict dense gatherings. At those times California had suffered 23 deaths and New York had accumulated 157. Eventually, the first wave peaked in California at 3.5 dpM/day in August; New York reached 38 dpM/day in mid-April.

A key factor in the COVID-19 development is the incredibly fast buildup of infections under normal-behavior conditions. As illustrated under Discussion, in New York state more than a million persons had been infected by the time the first person died. At this point any additional imported infectious persons are negligible compared to domestically induced infections.

Therefore, import restrictions can serve to limit the influx of infections to the degree that available test/track/quarantine resources can maintain control, or delay the onset. But they will be effective only if applied long before a significant number of deaths occur. Therefore, the COVID-19 pandemic on the U.S. west coast was delayed by restricting travel from China, but a restriction on travel from Europe would have been effective only if it were applied before Feb. 18, 2020.

### More or Less Social Restrictions at State/Local Level

Unwittingly the U.S. performed an experiment to evaluate different approaches to controlling the impact of the COVID-19 pandemic because the political controversy encouraged “Blue” states to be more aggressive than “Red” states. The resulting deaths is a measure of the relative value of the approaches. Unfortunately, data are not yet available on another important measure: the relative impact on the states’economies. There is no doubt that it was severe and that pre-emptive restrictions made it more so.

We compare the pandemic history in three states for insight: California and New York as solidly “Blue” states and Florida as a representative “Red” state. The final death tolls are 1570 dpM for California, 2115 dpM for New York and 1710 dpM for Florida. The first-wave death tolls were 109 dpM for California, 1189 for New York and 114 for Florida. In New York the first wave accounted for 56% of the eventual deaths; in California and Florida it accounted for about 7%.

In California Gov. Newsom imposed in March 2020 the earliest restrictions on large gatherings. In New York Gov. Cuomo followed suit, but the death rate had already soared. In Florida, Gov. DeSantis also restricted activities to essential services. During the summer and early fall of 2020, the ongoing relatively benign death rate caused Gov. Newsom and Gov. Cuomo to adjust restrictions, but in Sept. 2020 Gov. DeSantis nullified most restrictions imposed by local authorities. The resulting Dec./Jan. peak enabled California to reach death levels comparable to New York.

We must rate all three approaches as failures. The motivations were worthy: protect lives and protect the state’s economy. Neither goal was achieved. The only conclusions consistent with the data are:

> Pre-emptive restrictions are more effective than those applied when the death rate is already significant.
>
> The population tires of long-term restrictions and relaxes them, even while the death rate remains significant.

Instead, we study the two states that were relatively successful in controlling the pandemic: Hawaii and Alaska, one “Blue” and one “Red”. Their final deaths were comparable at 353 dpM and 495 dpM, respectively. Their remoteness enabled them to control ingress of infected persons, but the initial deaths in Alaska coincided with those in California; in Hawaii they occurred 9 days later. As we’ve argued above, at this stage home-grown infections outnumber imported ones. Alaska has the advantage of the lowest state population density at 1.3 persons/sq. mile, but Hawaii’s at 219 persons/sq. mile is comparable to California at 246 persons/sq. mile. The state governments provided guidance but did not impose severe restrictions other than controlling ingress.

Thus, we are forced to speculate further to explain the data. Without direct evidence to support it, we offer the following hypothesis:

> The remoteness of Hawaii, Alaska and New Zealand promotes in their citizens a sense of pride that induces them to exercise extra care to protect their state from pandemic infection. Restricting ingress from elsewhere is one manifestation of that pride. Conscientiously following guidance for social separation is another.

## Footnotes

* All such historical data were taken from Wikipedia summaries. They can be accessed on the Internet by a search for “COVID-19, Wiki,” with the name of state or nation.

† We must distinguish between two types of tests: virus and antivirus. A positive result on a virus test implies that there is active virus in that the person, i.e., he/she is infected and possibly infectious. A positive antivirus test establishes that the person has developed antibodies, but the test does not distinguish between active cases and those who have recovered from the infection. Hopefully, a positive antivirus result means that the person is immune from re-infection, at least for some time.

‡ CAUTION: This section combines objective data with context based on the author’s interpretation of contemporary news reports. While the implied subjectivity violates normal scientific standards, there should be no doubt that political context was a major causal factor in the development of the COVID-19 pandemic in the U.S. The reader is encouraged to apply his/her own interpretation to the data.

§ Author’s note: The data used to formulate these first three conclusions were available on the Internet by May 1, 2020.

** This is derived by assuming exponential growth for 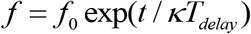 and approximating (1- *f*) ≈ 1.

†† The limiting case of Eq. 1 is readily solved by letting *f* = 1/*g*, yielding an easily solved linear equation for *g*: find 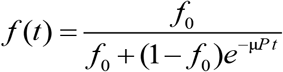, where *f*_o_ is a small initial seed value for *f*.

‡‡ We used an arbitrary seed value of *f*_*0*_ = 1*10^−8^.

